# From first to second wave: follow-up of the prospective Covid-19 cohort (KoCo19) in Munich (Germany)

**DOI:** 10.1101/2021.04.27.21256133

**Authors:** Katja Radon, Abhishek Bakuli, Peter Pütz, Ronan Le Gleut, Jessica Michelle Guggenbuehl Noller, Laura Olbrich, Elmar Saathoff, Mercè Garí, Yannik Schälte, Turid Frahnow, Roman Wölfel, Michael Pritsch, Camilla Rothe, Michel Pletschette, Raquel Rubio-Acero, Jessica Beyerl, Dafni Metaxa, Felix Forster, Verena Thiel, Noemi Castelletti, Friedrich Rieß, Maximilian N. Diefenbach, Günter Fröschl, Jan Bruger, Simon Winter, Jonathan Frese, Kerstin Puchinger, Isabel Brand, Inge Kroidl, Andreas Wieser, Michael Hoelscher, Jan Hasenauer, Christiane Fuchs, on behalf of the KoCo19 study group

## Abstract

**Background:** In the 2^nd^ year of the Covid-19 pandemic, knowledge about the dynamics of the infection in the general population is still limited. Such information is essential for health planners, as many of those infected show no or only mild symptoms and thus, escape the surveillance system. We therefore aimed to describe the course of the pandemic in the Munich general population living in private households from April 2020 to January 2021.

**Methods:** The KoCo19 baseline study took place from April to June 2020 including 5313 participants (age 14 years and above). From November 2020 to January 2021, we could again measure SARS-CoV-2 antibody status in 4,433 of the baseline participants (response 83%). Participants were offered a self-sampling kit to take a capillary blood sample (dry blood spot; DBS). Blood was analysed using the Elecsys**®** Anti-SARS-CoV-2 assay (Roche). Questionnaire information on socio-demographics and potential risk factors assessed at baseline was available for all participants. In addition, follow-up information on health-risk taking behaviour and number of personal contacts outside the household (N=2768) as well as leisure time activities (N=1263) were collected in summer 2020.

**Results:** Weighted and adjusted (for specificity and sensitivity) SARS-CoV-2 sero-prevalence at follow-up was 3.6% (95% CI 2.9-4.3%) as compared to 1.8% (95% CI 1.3-3.4%) at baseline. 91% of those tested positive at baseline were also antibody-positive at follow-up. While sero-prevalence increased from early November 2021 to January 2021, no indication of geospatial clustering across the city of Munich was found, although cases clustered within households. Taking baseline result and time to follow-up into account, men and participants in the age group 20-34 years were at the highest risk of sero-positivity. In the sensitivity analyses, differences in health-risk taking behaviour, number of personal contacts and leisure time activities partly explained these differences.

**Conclusion:** The number of citizens in Munich with SARS-CoV-2 antibodies was still below 5% during the 2^nd^ wave of the pandemic. Antibodies remained present in the majority of baseline participants. Besides age and sex, potentially confounded by differences in behaviour, no major risk factors could be identified. Non-pharmaceutical public health measures are thus still important.

## Introduction

The SARS-CoV-2 virus affected almost all nations within a few weeks. Given the nature of the virus, with a large proportion of infected individuals infected present only mild symptoms or no symptoms at all. Therefore, population-based sero-prevalence studies are necessary to estimate the true prevalence of the infection in the population. Starting in March 2020, such sero-prevalence studies have been conducted in many countries, mostly during or after the first wave of the pandemic (1). Depending on the serological test used, the type of sample drawn, the timing of the study, and the region, general population sero-prevalence ranged from <0.1% in Brazil to well over 20% in the USA (2). For the German context, we reported a sero-prevalence of 1.8% in Munich, sampled towards the end of the first wave in Germany (3).

Following the introduction of public health measures (lock-down including school closures) in March 2020 in Germany, the first wave of the pandemic was perceived as relatively mild with around 6,000 cases registered in Munich during this period (Munich population ∼1.5 Mio). Between June and October, public health measures were reduced, although physical distancing of 1.5 m between two persons, avoidance of mass events, and obligatory use of face masks, e.g. in restaurants and shops, were still required. Subsequently, officially registered monthly case numbers in Munich rose from 389 in June to 7,181 in October 2020. A partial national lock-down was implemented on November 2^nd^, 2020. After a further rise in officially registered case numbers and COVID-19 related deaths (Figure 1), national lock-down measures were increased from December 16^th^, 2020 on, including closure of schools, shops (other than grocery and drug stores), restaurants, and hotels.

**Figure 1:**
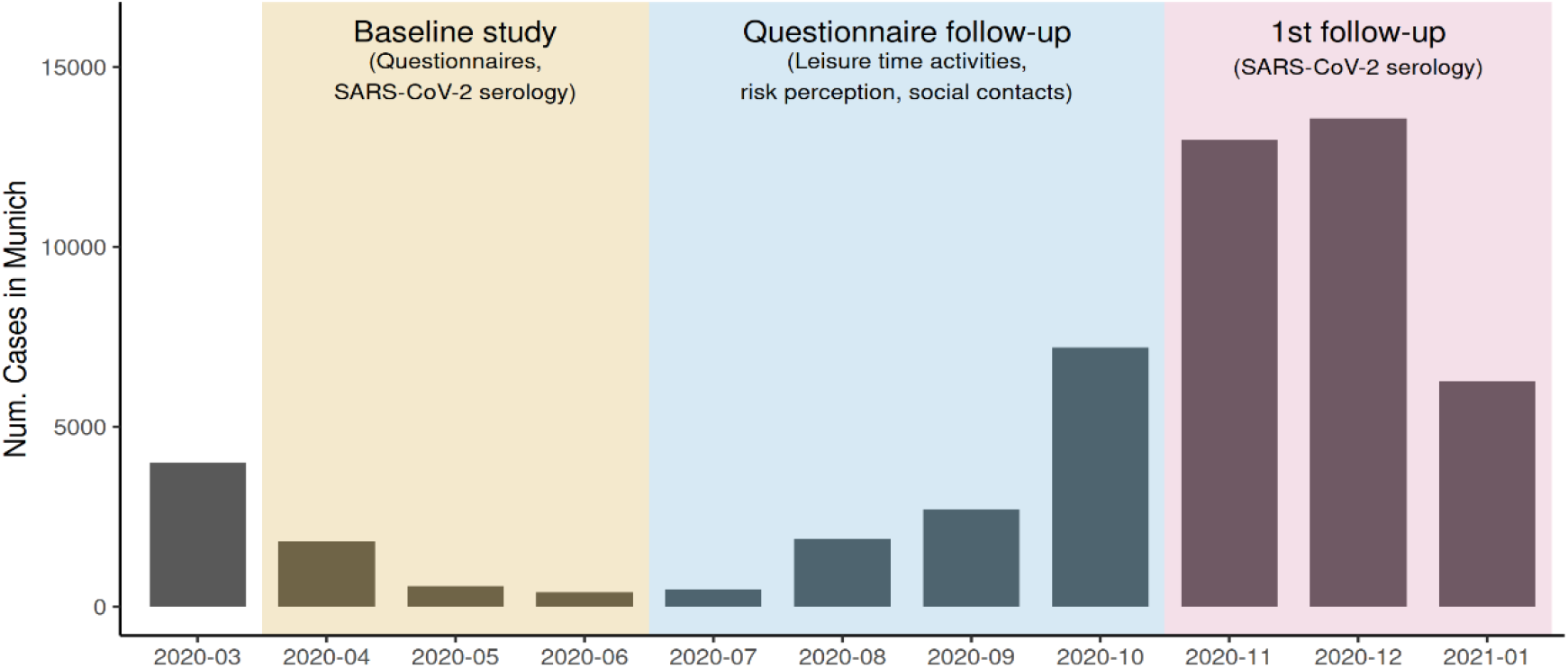
Monthly number of registered SARS-CoV-2 infections in Munich and course of the KoCo19 study (case numbers taken from: https://corona.stat.uni-muenchen.de/nowcast/)

Given that asymptomatic and mildly symptomatic cases escape surveillance systems, prospective population-based cohort studies offer the chance to better understand the course of disease in the general population. They are independent of testing strategies and help to identify the population at risk over time. In addition, they provide an indication of population groups less well protected by public health measures. We therefore followed up the participants of the Munich COVID-19 cohort (KoCo19). Besides the 1^st^ antibody follow-up, we assessed behavioural factors by online questionnaires. The baseline study took place from April to June 2020, the questionnaire follow-up in summer 2020 and the 1^st^ antibody follow-up was realised from early November 2020 to January 2021 (Figure 1). On December 1^st^ 2020 the KoCo19 cohort joined the ORCHESTRA (Connecting European Cohorts to Increase Common and Effective Response to SARS-CoV-2 Pandemic) project.

## Methods

### Study population and field work

#### Baseline SARS-CoV-2 antibody and questionnaire study

We described the baseline study in detail in (4). In short, a random sample of the Munich population living in private households was drawn by random walk method. All household members older than 13 years were invited to provide a serum sample and to answer an online questionnaire. Serum samples were analysed for SARS-CoV-2 antibodies using the Elecsys**®** Anti-SARS-CoV-2 (Roche) test (5). Field work for the baseline study took place between April 5^th^ and June 12^th^, 2020.

#### Questionnaire follow-up

An online questionnaire covering risk behaviour, health related items, and psychosocial aspects (hereafter “behaviour questionnaire”) was offered from June 4^th^ to October 31^st^, 2020 to all 5,240 participants who did not withdraw from the study. In parallel, an online-questionnaire on leisure time behaviour was available (hereafter “leisure time questionnaire”). We split the questionnaire into two, because long questionnaires are less likely to be completed (6). Participants recruited in April (May to June) 2020 received an invitation via e-mail on June 4^th^ (June 25^th^) with subsequent reminders and telephone follow-ups. In total, 3,400 participants completed the behaviour questionnaire and 1,390 participants the leisure time questionnaire.

#### 1^st^ SARS-CoV-2 antibody follow-up

On November 2^nd^ 2020, we started the 1^st^ antibody follow-up by sending out boxes with a self-sampling kit to take a capillary blood samples (dry blood spot; DBS) to the 5,292 participants (2,978 households) of the baseline study (Figure 1). Between baseline and follow-up, 77 participants withdrew from the study and were thus not contacted for the follow-up. Instructions for self-sampling were provided, including a video tutorial (https://www.youtube.com/watch?v=vpZUzuQV10E&feature=emb_title). Samples were collected using a barcode-labelled neonatal screening filter card (Euroimmun ZV 9701-0101) with circles indicating where the blood should be collected. Afterwards, participants should dry the filter card at least 12 hours at room temperature, pack them in the sealable plastic pouch, place the plastic pouch into the prepaid envelope, and ship the envelope by mail to the laboratory. In case of handling difficulties, our telephone and e-mail hotline were available for any questions.

From November 2^nd^ to January 31^st^, 2021, we received 4,444 DBS samples from 2,571 households (individual response 84%, household response 86%). Participants not being able to collect a DBS on their own (N=29) and those with intermediate results (N=34, s. laboratory methods) were offered a full-blood test at our centre. For the latter group, this served to clarify the DBS result. However, 11 of the 34 participants with intermediate results in the DBS did not show up at our centre and thus had to be excluded from analyses, leaving 4,433 subjects with baseline questionnaire, baseline serology and follow-up DBS data for the main analyses (Figure 2).

**Figure 2:**
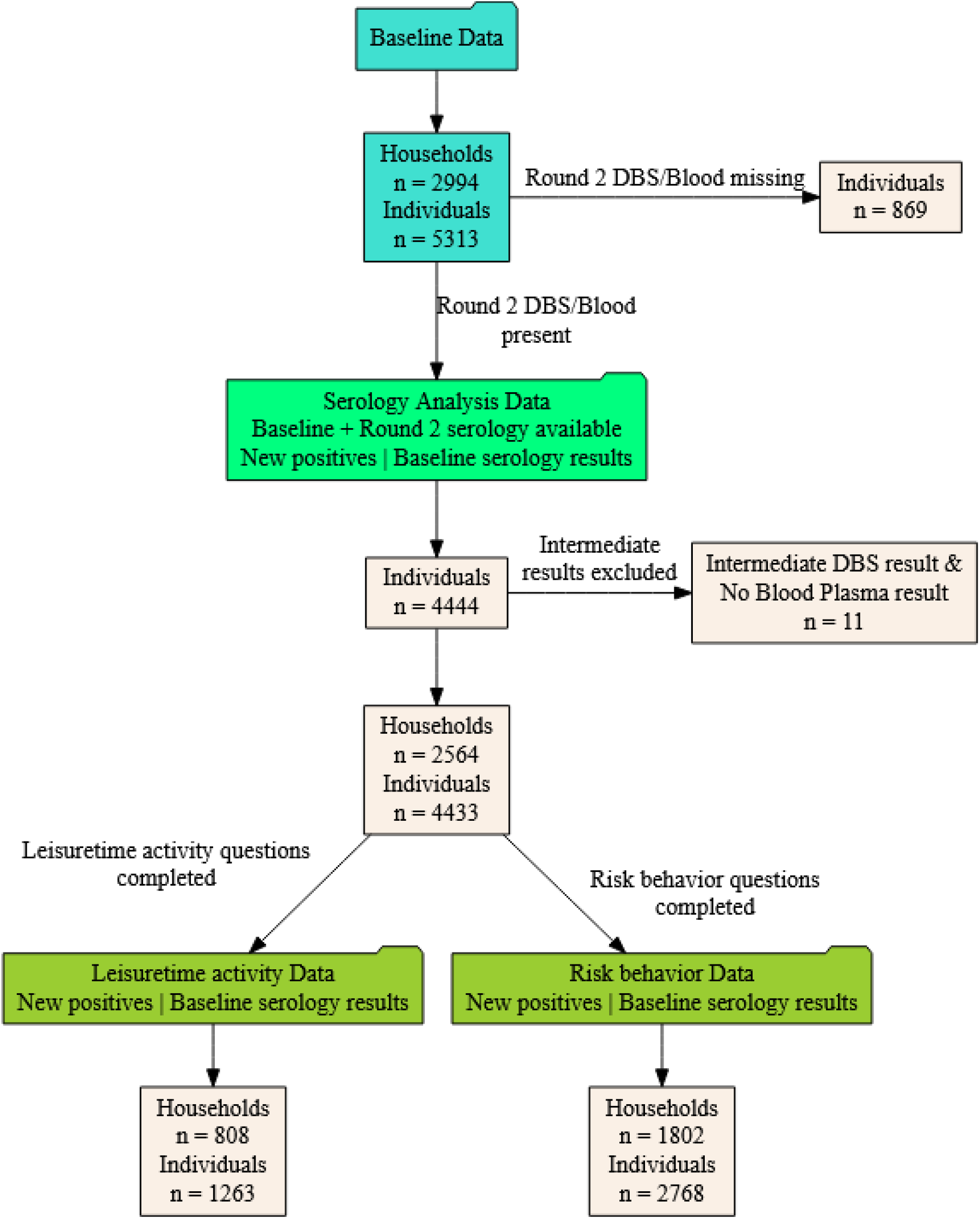
Flow chart of obtaining the study population

### Questionnaire data

The following items were considered for the analyses presented in this paper: Baseline individual questionnaire:

- Socio-demographics: age, sex (male, female), schooling (<12 years, ≥12 years, in school), current job (employed, self-employed, not working (unemployed, retired, parental leave, sabbatical, students), others (voluntary social year, military service, part-time jobber, reduced working hours))
- Country of birth: Germany, others
- Smoking: current, ex, never smokers
- Chronic conditions: diabetes, cardiovascular diseases, autoimmune diseases, respiratory diseases (yes vs no)

Baseline household questionnaire:

- Household size: 1, 2, 3-4, >5 inhabitants
- Household income: ≤2,500 €, 2,500-≤4,000 €, 4,000-≤6,000 €, >6000 €
- Living area per inhabitant: ≤30 sqm, 30-≤40 sqm, 40-≤55 sqm, >55 sqm
- Household type: single, couple, family, others (shared apartments by e.g., students, subleasing, and assisted accommodation)
- Housing type: building with 1-2 apartments, 3-4 apartments, ≥ 5 apartments

Follow-up questionnaire:

- Self-estimated health-related risk taking behaviour (10-level Likert scale from “not at all risk tolerant” to “very risk tolerant”): Dichotomised into not high (≤5, Quartile 3) and high self-estimated health-related risk taking behaviour (>5)
- Personal contacts: Five questions on places of personal contacts outside the own household during the two weeks before answering the questionnaire (meeting people, grocery shopping, shopping, use of public transport, work outside home), each assessed on a 5-level Likert scale: not at all (=1); once per week (=2); 2-4 times per week (=3); 5 times per week (=4); more often (=5). Places of personal contacts were multiplied by frequency of contacts (0 contacts (=0), 1 contact (=1), 2-4 contacts (=2) and 5+ contacts (=3)) and summed up, resulting in a score ranging from 0 to 25. The score was dichotomised into lower number of personal contacts (≤8, Median) and higher number of personal contacts (>8).
- Number and intensity of leisure time activities before the pandemic (in February 2020): For that time, 16 activities assessed on a 5-level Likert scale from “never” (=0) to “very often” (=4): visit family member; visit friends; going out with friends; attend a party, festival, bar, pub or disco; go to the cinema; attend a theatre, opera or ballet performance; work out in a gym; visit a swimming pool; visit a sauna; skiing; train for a team sport or take part in sporting competitions; watch a sports game or event live outdoors; watch a sports game or event live indoors; worship attendance; play an instrument in an orchestra; sing in a choir. Activities were multiplied by the Likert scores and summed up resulting in a score from 0 to 64. The score was dichotomised into non-high leisure time activities (≤11, Quartile 3) and high leisure time activities (>11).
- Number and intensity of leisure time activities two weeks prior to the follow-up questionnaire: The score for leisure time activities at follow-up was built the same way as the score for leisure time activities before the pandemic. However, the number of leisure time activities was only seven at that time as many activities were not possible due to the restrictions related to the pandemic: visit family member; visit friends; going out with friends; visit a swimming pool; worship attendance; play an instrument in an orchestra; sing in a choir. Therefore, the resulting score only ranged from 0 to 28. The score was dichotomised into non-high leisure time activities at follow-up (≤5, Quartile 3) and high leisure time activities (>5).

### Laboratory method and cross-validation with blood samples

Filter paper cards were further processed if at least two of the five circles on the card were completely soaked with blood. Valid samples were stored at 4 °C until analysis. Before analysis, filter paper cards were equilibrated to room temperature and three blood-soaked smaller circles (diameter 3.2 mm) of each filter paper card were automatically punched into a 96-wells plate (Panthera-Puncher(tm) 9, PerkinElmer). After elution, samples were transferred to a Cobas e801 module (Roche) compatible sample micro cup (Roche, 05085713001) for analysis using the Elecsys**®** Anti-SARS-CoV-2 assay (Roche). Based on our validation study, DBS samples were considered positive if SARS-CoV-2 antibody levels were ≥0.12. Samples with SARS-CoV-2 antibody levels in the range between 0.09 and 0.12 were considered intermediate, and subsequently confirmed by plasma samples (s. Study population and field work). All other samples were considered negative. Compared to full blood samples, sensitivity of the DBS method was 99.2% and specificity 98.7%. Details of the laboratory methods are described in (7).

### Statistical analyses

All statistical analyses were performed using the statistical software R (version 4.0.3, R Development Core Team, 2020).

The SARS-CoV-2 sero-prevalence was estimated primarily based on the DBS test results of the study participants applying the classification as described above (Laboratory methods). If the DBS test yielded an intermediate result, we considered the result of the full blood sampling. As described in (5), an optimised cut-off of 0.4218 for the full blood sampling was used to predict SARS-CoV-2 sero-positivity with an estimated specificity and sensitivity of 99.7% and 88.6%, respectively (with regard to PCR test results considered as ground truth). We used these estimates to adjust the prevalence for the imperfect test performance (8). The specificity and sensitivity of DBS with regard to full blood samples being very high, additional adjustment was omitted (Appendix Text and Table S1).

The prevalence (adjusted or unadjusted for the specificity and the sensitivity of the test) was calculated in two different ways: including the information from the sampling design of the cohort (3) via the use of a weighting scheme, or without it. To account for the sampling design, the sampling weights computed at baseline (inverse of the probability of each individual to be included in the sample) were used for the follow-up analysis. These sampling weights were corrected for the attrition observed at follow-up by modelling the underlying non-response mechanism and estimating probabilities of response for each unit. Ten response homogeneity groups (where we assumed the non-response to be completely at random, (9)) were created using the deciles of the estimated probabilities of response. These weights adjusted for the non-response were calibrated (10) on updated information from the Munich population (at 31.12.2020) in order to mirror the age, sex, country of birth, presence of children in the household and single member household structures. Moreover, to correct the sample for the loss of positive cases at follow-up, the sampling weights were calibrated on the estimated number of positive cases at baseline. Weighted prevalence estimates were calculated using these calibrated weights, and the associated 95% confidence intervals were computed based on variance estimators based on linearization (10) and residual (10, 11) techniques. These variance estimates were computed in order to account for every step in the selection process of the units, i.e., V = V1 + V2 with V1 the variance due to the sampling design and V2 the one due to the non-response (12). For unweighted prevalence estimates, confidence intervals were determined by using a nonparametric cluster bootstrap procedure that accounts for household clustering (13). To that end, 5,000 bootstrap datasets were generated each by sampling nh households with replacement from the original sample of nh households. The sero-prevalence was estimated in each bootstrap sample and the 2.5 and 97.5 percentiles of the resulting 5,000 estimates defined the 95% confidence intervals.

To analyse spatial clustering, we considered the mean within-cluster variance of the binary test results, with cluster variables being households, buildings, and geospatial clusters of different sizes. We performed a non-parametric approximate permutation test with 10,000 random permutations of cluster assignments. To account for household clustering, only full households were permuted when considering buildings and geospatial clusters (14). In addition to this, we performed borough level sero-prevalence mapping using Conditional Auto Regressive Models which account for the spatial autocorrelation among neighbouring boroughs by using random effects. This allowed us to investigate if sero-prevalence was associated with the population density or not, as well as obtaining Borough/District level estimates within the city of Munich (Appendix 2) (15-18).

We used generalised linear mixed models (GLMMs using the logit link function) to analyse the association between potential risk factors and SARS-CoV-2 sero-positivity at 1^st^ follow-up, with a random effect for households to account for within household clustering of the data. Odds Ratio estimates and the corresponding confidence intervals were obtained applying a Bayesian framework with uniform priors on the regression estimates using the brms (Bayesian Regression Models using ‘Stan’) package in R (19, 20). To account for missing data in covariates, we used the Joint Analysis and Imputation of Incomplete Data Framework (JointAI) in R for sensitivity analyses (21, 22). In these sensitivity analyses, broad normal priors with mean zero and standard deviation 100 were used. The regression estimates were adjusted for the SARS-CoV-2 serology results at baseline, the time elapsed since baseline visit, age, and sex of the individual. Essentially, this adjustment for baseline positivity allowed us to obtain risk factors associated with newly incident cases within our cohort over and above the baseline positives.

To explore the importance of behavioural factors and leisure-time activities for the incidence of infection between baseline and follow-up, we used data of the 1^st^ questionnaire follow-up combined with the DBS results. For these analyses, we included information of 2,768 participants who responded to the behaviour questionnaire and had serology results; for the leisure time activities, we had questionnaire information for 1,263 persons with serological results. Due to the large proportion of missing questionnaire data, we restricted these analyses to complete data and aggregated at the levels of the outcome variables. We analysed the incidence of new SARS-CoV-2 infections (as binomial outcome for proportions) between baseline and follow-up stratified for risk behaviour, leisure time activities, sex and age, using the count of new positives among the observed. Similar models were also applied to evaluate the association of the population densities at the constituency level and the trend in sero-prevalence estimates using aggregated data.

## Results

Follow-up participants compared to participants lost to follow-up were more likely to be between 35 and 79 years old, born in Germany (84% vs. 74%), and to have a higher socio-economic status. The latter was indicated, e.g., by level of education, household income, living area, and type of building. In addition, the sero-prevalence of SARS-CoV-2 at baseline was lower among follow-up participants (1.6%) compared to baseline only participants (2.6%; Table 1). These losses of positive cases at follow-up led to an underestimation of the total number of people tested positive at baseline in Munich (22,064 vs. 25,900 using all participants at baseline). To correct for this attrition bias, the weights at follow-up were calibrated on the estimate of positive cases at baseline, in addition to the other margins used to mirror the Munich structure.

**Table 1:**
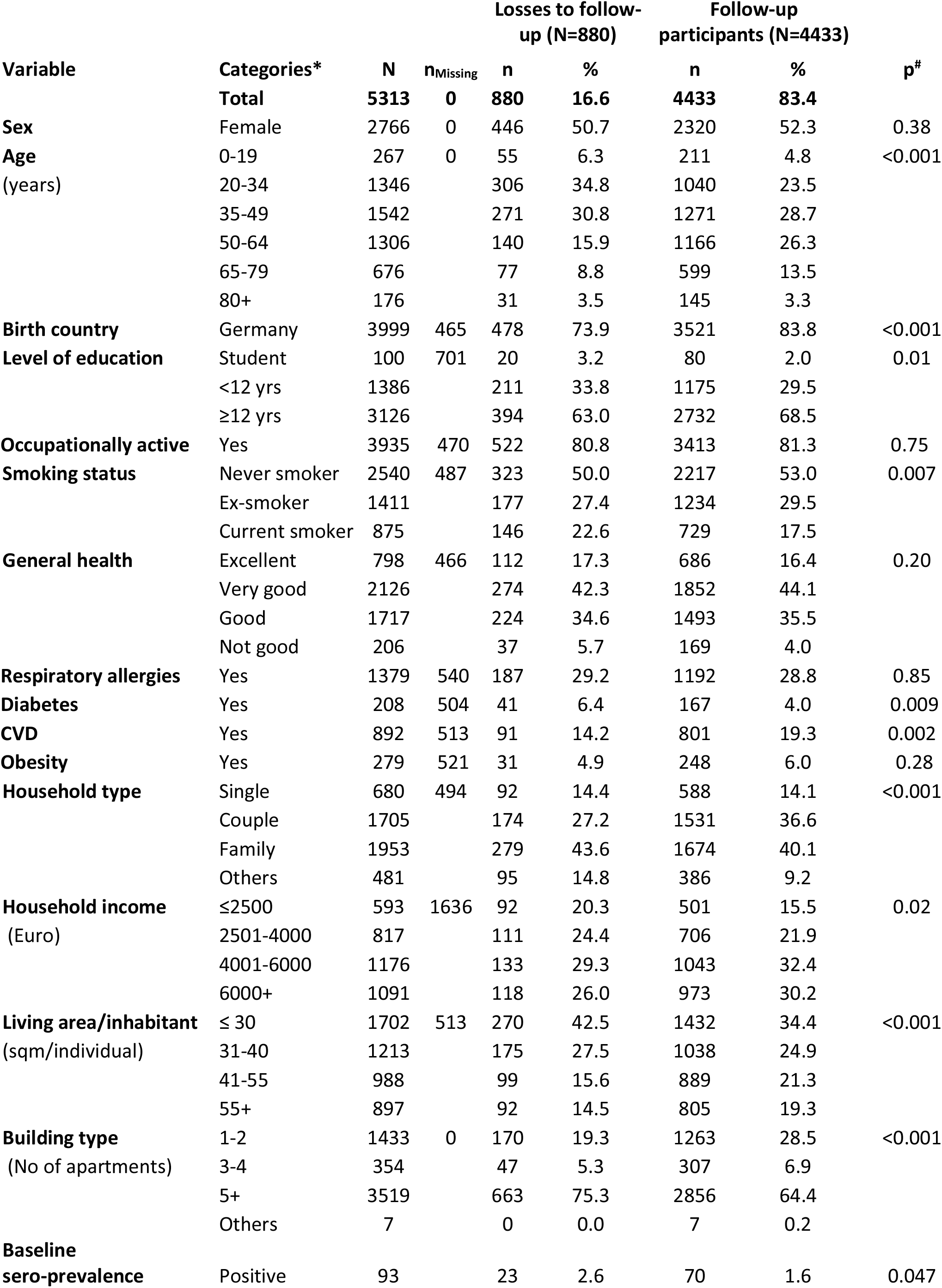
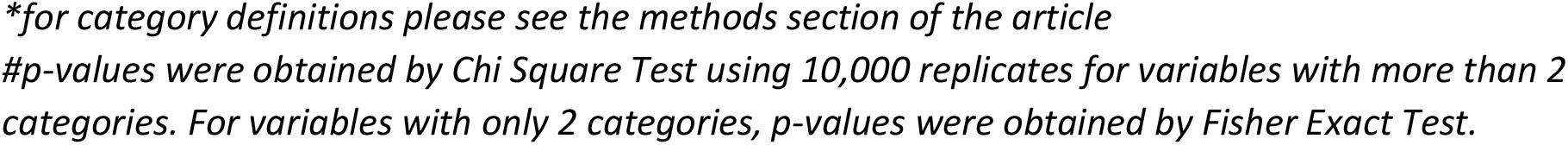
Descriptive data of the KoCo-19 follow-up participants in comparison to participants only taking part in the baseline study (“Losses to follow-up”)

| Variable | Categories* | N | n<sub>Missing</sub> | Losses to follow-up (N=880) |  | Follow-up participants (N=4433) |  | p<sup>#</sup> |
| --- | --- | --- | --- | --- | --- | --- | --- | --- |
|  |  |  |  | n | % | n | % |  |
|  | **Total** | **5313** | **0** | **880** | **16.6** | **4433** | **83.4** |  |
| **Sex** | Female | 2766 | 0 | 446 | 50.7 | 2320 | 52.3 | 0.38 |
| **Age**<br>(years) | 0-19 | 267 | 0 | 55 | 6.3 | 211 | 4.8 | <0.001 |
|  | 20-34 | 1346 |  | 306 | 34.8 | 1040 | 23.5 |  |
|  | 35-49 | 1542 |  | 271 | 30.8 | 1271 | 28.7 |  |
|  | 50-64 | 1306 |  | 140 | 15.9 | 1166 | 26.3 |  |
|  | 65-79 | 676 |  | 77 | 8.8 | 599 | 13.5 |  |
|  | 80+ | 176 |  | 31 | 3.5 | 145 | 3.3 |  |
| **Birth country** | Germany | 3999 | 465 | 478 | 73.9 | 3521 | 83.8 | <0.001 |
| **Level of education** | Student | 100 | 701 | 20 | 3.2 | 80 | 2.0 | 0.01 |
|  | <12 yrs | 1386 |  | 211 | 33.8 | 1175 | 29.5 |  |
|  | ≥12 yrs | 3126 |  | 394 | 63.0 | 2732 | 68.5 |  |
| **Occupationally active** | Yes | 3935 | 470 | 522 | 80.8 | 3413 | 81.3 | 0.75 |
| **Smoking status** | Never smoker | 2540 | 487 | 323 | 50.0 | 2217 | 53.0 | 0.007 |
|  | Ex-smoker | 1411 |  | 177 | 27.4 | 1234 | 29.5 |  |
|  | Current smoker | 875 |  | 146 | 22.6 | 729 | 17.5 |  |
| **General health** | Excellent | 798 | 466 | 112 | 17.3 | 686 | 16.4 | 0.20 |
|  | Very good | 2126 |  | 274 | 42.3 | 1852 | 44.1 |  |
|  | Good | 1717 |  | 224 | 34.6 | 1493 | 35.5 |  |
|  | Not good | 206 |  | 37 | 5.7 | 169 | 4.0 |  |
| **Respiratory allergies** | Yes | 1379 | 540 | 187 | 29.2 | 1192 | 28.8 | 0.85 |
| **Diabetes** | Yes | 208 | 504 | 41 | 6.4 | 167 | 4.0 | 0.009 |
| **CVD** | Yes | 892 | 513 | 91 | 14.2 | 801 | 19.3 | 0.002 |
| **Obesity** | Yes | 279 | 521 | 31 | 4.9 | 248 | 6.0 | 0.28 |
| **Household type** | Single | 680 | 494 | 92 | 14.4 | 588 | 14.1 | <0.001 |
|  | Couple | 1705 |  | 174 | 27.2 | 1531 | 36.6 |  |
|  | Family | 1953 |  | 279 | 43.6 | 1674 | 40.1 |  |
|  | Others | 481 |  | 95 | 14.8 | 386 | 9.2 |  |
| **Household income**<br>(Euro) | ≤2500 | 593 | 1636 | 92 | 20.3 | 501 | 15.5 | 0.02 |
|  | 2501-4000 | 817 |  | 111 | 24.4 | 706 | 21.9 |  |
|  | 4001-6000 | 1176 |  | 133 | 29.3 | 1043 | 32.4 |  |
|  | 6000+ | 1091 |  | 118 | 26.0 | 973 | 30.2 |  |
| **Living area/inhabitant**<br>(sqm/individual) | ≤ 30 | 1702 | 513 | 270 | 42.5 | 1432 | 34.4 | <0.001 |
|  | 31-40 | 1213 |  | 175 | 27.5 | 1038 | 24.9 |  |
|  | 41-55 | 988 |  | 99 | 15.6 | 889 | 21.3 |  |
|  | 55+ | 897 |  | 92 | 14.5 | 805 | 19.3 |  |
| **Building type**<br>(No of apartments) | 1-2 | 1433 | 0 | 170 | 19.3 | 1263 | 28.5 | <0.001 |
|  | 3-4 | 354 |  | 47 | 5.3 | 307 | 6.9 |  |
|  | 5+ | 3519 |  | 663 | 75.3 | 2856 | 64.4 |  |
|  | Others | 7 |  | 0 | 0.0 | 7 | 0.2 |  |
| **Baseline sero-prevalence** | Positive | 93 |  | 23 | 2.6 | 70 | 1.6 | 0.047 |

*\*for category definitions please see the methods section of the article*
*#p-values were obtained by Chi Square Test using 10,000 replicates for variables with more than 2 categories. For variables with only 2 categories, p-values were obtained by Fisher Exact Test.*

We received roughly half of the DBS samples (2,372 of 4,433; 54%) within 1 week of mailing (November 2^nd^ to November 8^th^). By week 2 (November 9^th^ to November 15^th^), more than three quarters were received (3,369 of 4,433; 76%). Most of the remaining samples were turned in between week 3 (N=372 from November 16^th^ to November 22^th^) and week 4 (N=343; November 23^rd^ to November 29^th^). Few samples were received in December 2020 and January 2021 (N=326; 7%).

The overall weighted and adjusted (for specificity and sensitivity) SARS-CoV-2 sero-prevalence at follow-up was 3.6% (95% CI 2.9-4.3%; Figure 3). The overall unweighted and adjusted sero-prevalence was 3.1% (95% CI 2.5-3.8%), increasing from 2.5% (95% CI 1.7-3.3%) in the first week of November to 4.0% (95% CI 1.6-6.8%) in the last week of November (Figure 4). About half of the participants with intermediate result in the DBS test had a positive test result when considering the plasma sample. As plasma samples were collected in December and January, the prevalence estimates in the latest weeks were artificially high. Yet, the overall upward trend remained after excluding the participants with intermediate DBS result (Appendix Figure S1, Figure S2).

**Figure 3:**
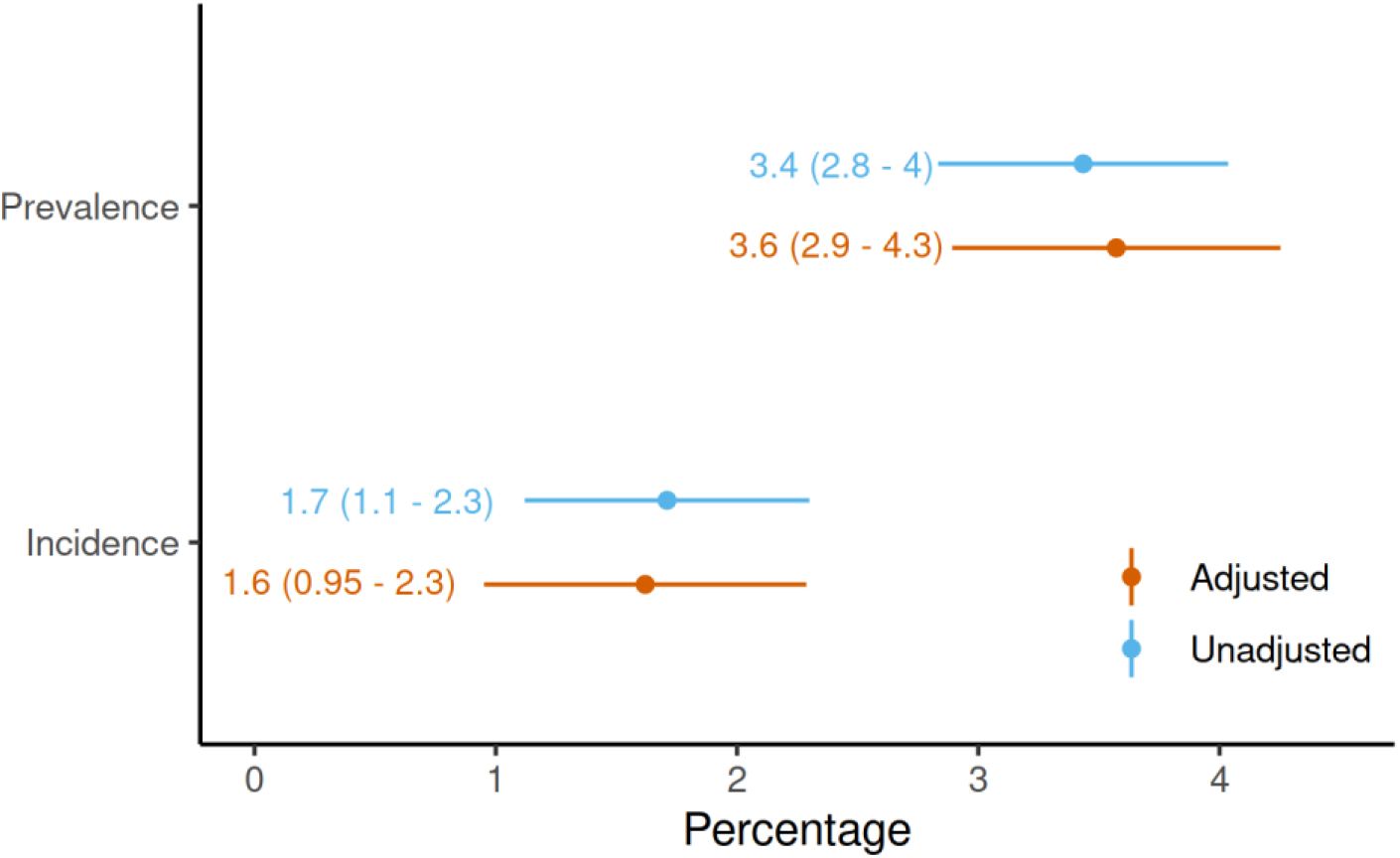
Weighted sero-prevalence and sero-incidence at follow-up adjusted (orange) and unadjusted (blue) for test specificity and sensitivity.

**Figure 4:**
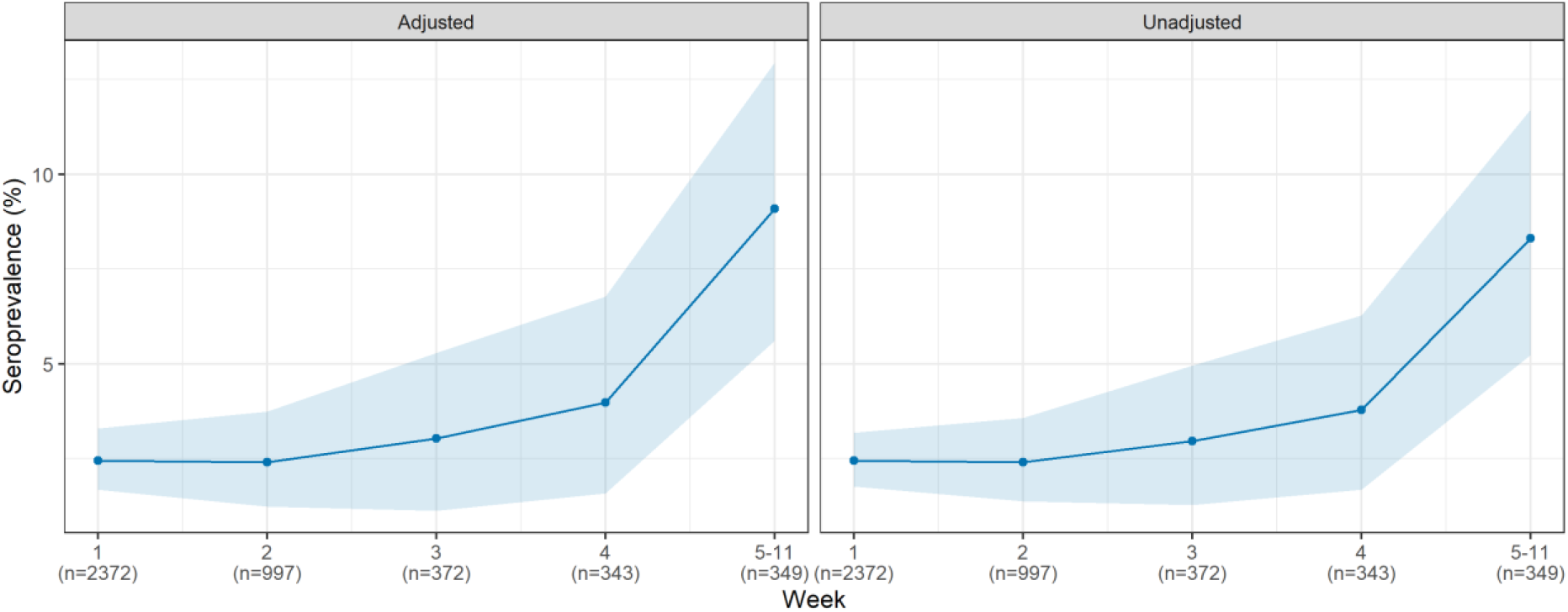
For sensitivity and specificity adjusted (left) and unadjusted (right) SARS-CoV-2 sero-prevalence over the follow-up period. The 95% confidence intervals for the weekly sero-prevalence were based on the 2.5 and 97.5 percentiles from 5,000 repetitions of a cluster bootstrap that accounts for within household clustering. The estimates do not account for sample weights. The estimation without accounting for within-household clustering but considering sample weights produced similar trends (Appendix Figure S1).

Most participants who were SARS-CoV-2 sero-positive at baseline continued to be sero-positive at follow-up (64 out of 70 sero-positive subjects at baseline). The weighted and adjusted SARS-CoV-2 sero-incidence (negative at baseline, positive at follow-up) was estimated at 1.6% (95% CI 0.95-2.3%) (Figure 3).

Looking at the geospatial distribution of SARS-CoV-2 sero-prevalence by Munich city boroughs (Figure 5), an increase was visible from the South-East to the North-West of the city, although these differences were rather small. The estimates of Moran’s I for spatial autocorrelation was 0.015 using the continuous distance based spatial neighbourhood matrix resulting in a p-value of 0.304, and hence was not statistically significant. Using a binary spatial neighbourhood matrix, the estimated Moran’s I was 0.025 with a corresponding p-value of 0.269 - thereby the conclusion remained unchanged. We also took population density as a potential risk factor into account and could not find any statistically significant association between population density in the constituency and SARS-CoV-2 sero-prevalence (Appendix Figure S3).

**Figure 5:**
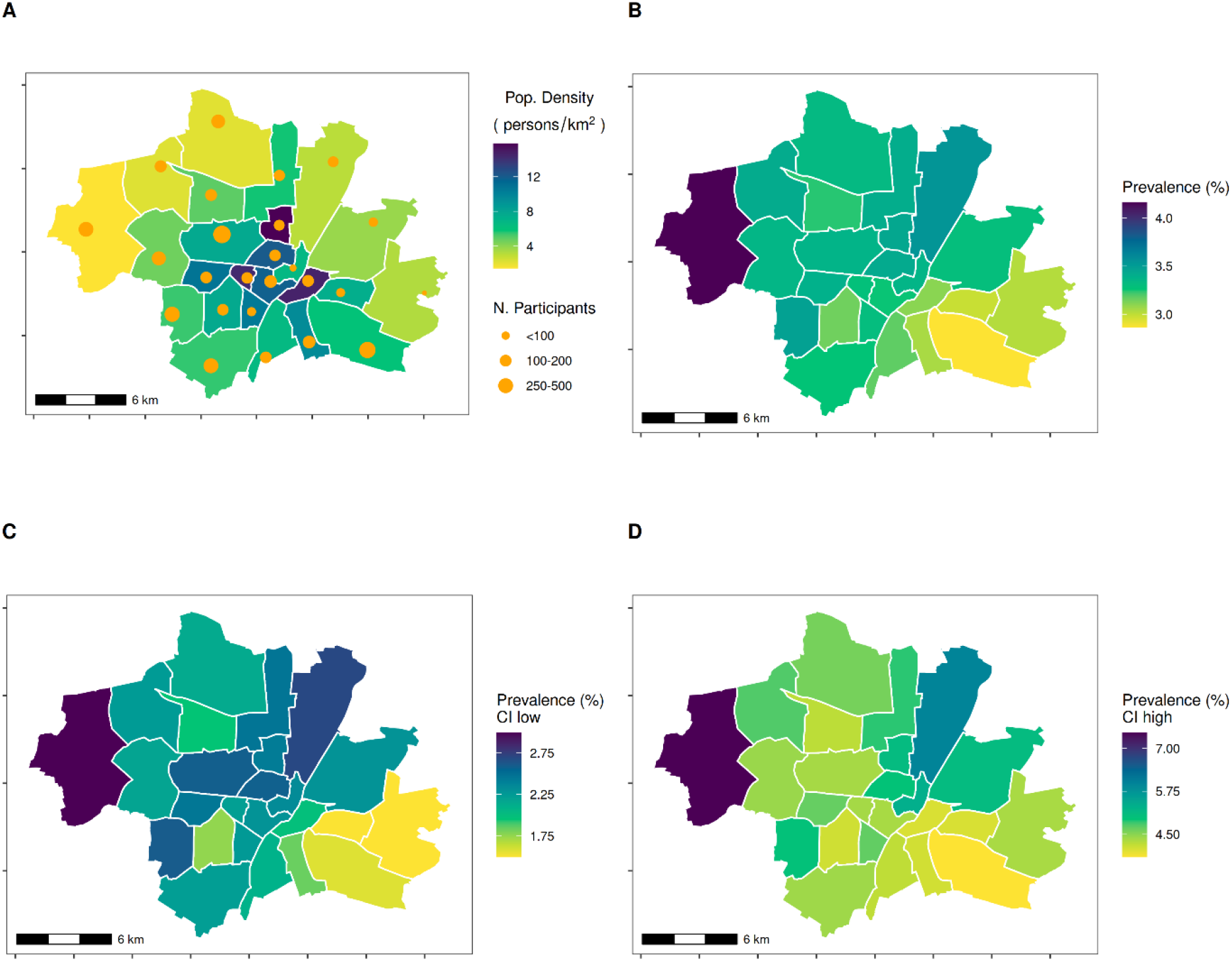
Geospatial distribution of the crude SARS-CoV-2 sero-positivity across boroughs in Munich. A Population density and number of participants in each city borough; B Weighted sample based SARS-CoV-2 sero-prevalence; C Lower 95% confidence bounds of the weighted SARS-CoV-2 sero-prevalence; D Upper 95% confidence bounds of the weighted SARS-CoV-2 sero-prevalence. The population density was taken from https://simple.wikipedia.org/wiki/Boroughs_of_Munich

The distribution of SARS-CoV-2 sero-positivity by covariates is shown in the Appendix (Table S3). Taking household clustering, time elapsed between baseline and follow-up, and baseline result into account, men had statistically significantly higher odds of sero-positivity at follow-up (OR adjusted for age: 2.4; 95% CI 1.0-6.0; Figure 6). In addition, SARS-CoV-2 sero-prevalence decreased with increasing age group. It was lower in participants living in small apartment houses compared to participants living in single houses (OR adjusted for age and sex 0.0002; 95% CI 0.0-0.14).

**Figure 6:**
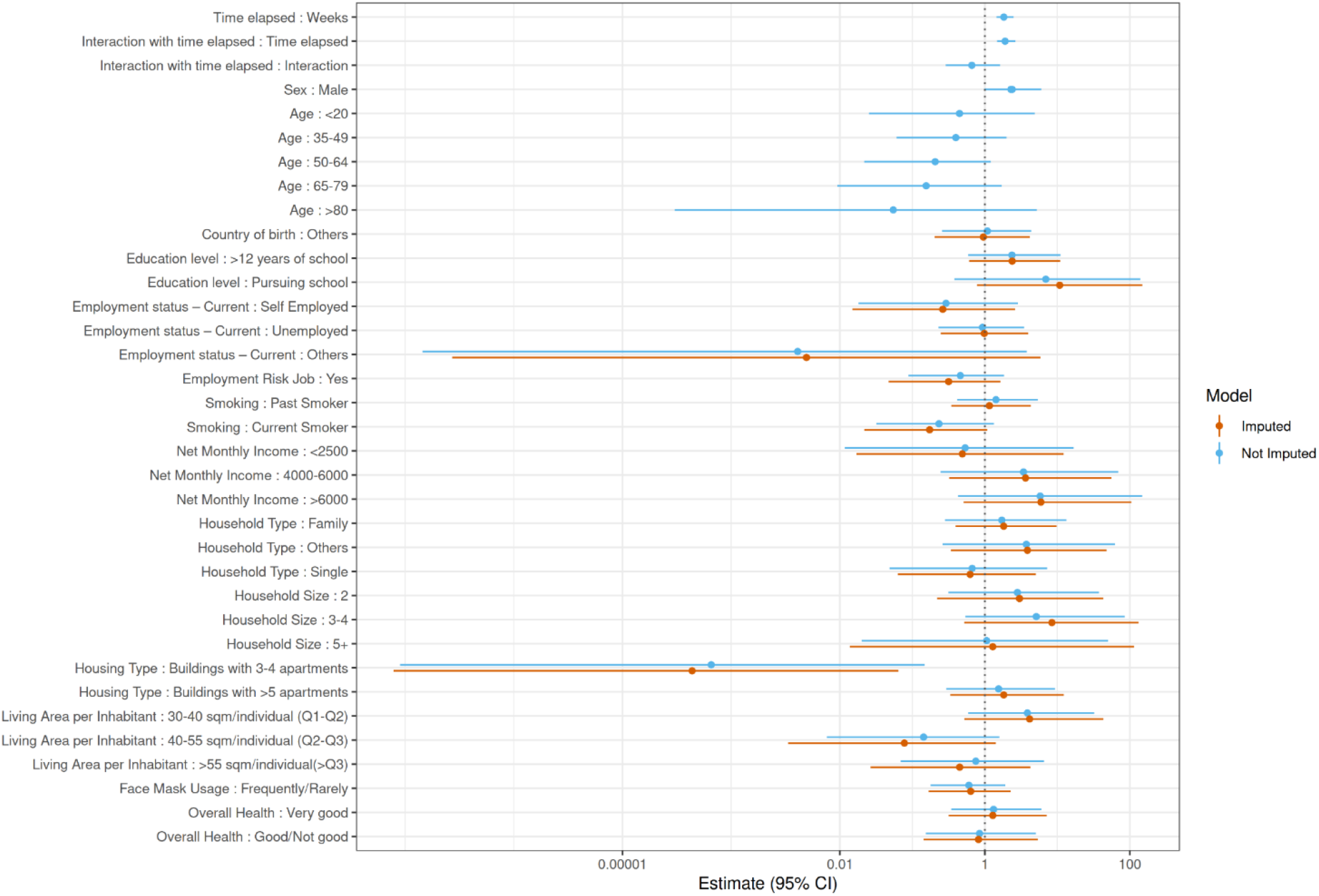
Association between potential risk factors and SARS-CoV-2 sero-positivity taking into account time between 1^st^ and 2^nd^ sampling, baseline result, age and sex. Unimputed (blue) and imputed (orange) GLM Models (Bayesian analysis).

The analysis of potential household and neighbourhood clustering indicated a highly significant within-household clustering, while no indications for neighbourhood transmission were observed (Figure 7, Appendix Figure S5).

**Figure 7:**
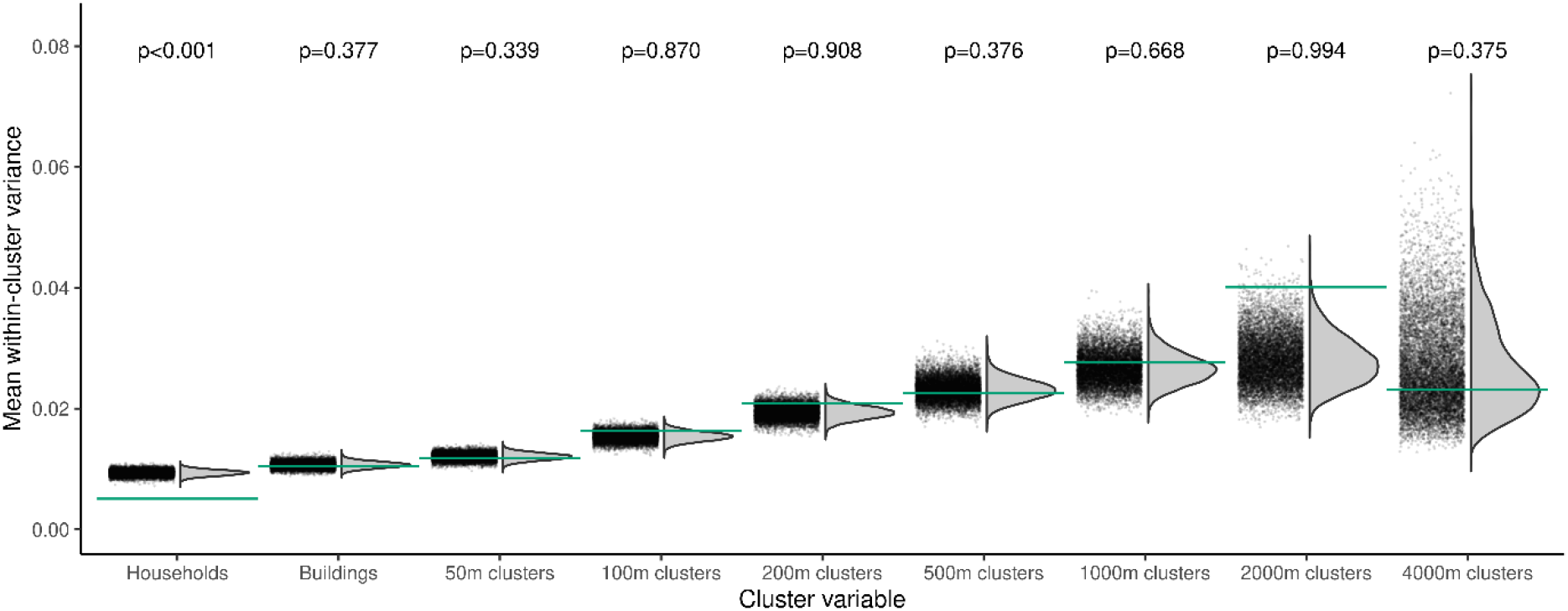
Proximity clustering of test outcomes at 2^nd^ sampling. The grey points and curves show the distribution of mean within-cluster variances for 10,000 random permutations of cluster assignments, the horizontal lines show the observed values. Cluster variables are households, buildings, and geospatial clusters of different sizes. Household membership was left invariant when considering buildings and geospatial clusters. P-values indicate the one-sided probability of observing smaller than observed values under random cluster assignments.

### Sensitivity analyses on behavioural factors and SARS-CoV-2 sero-incidence

In order to identify which behavioural factors might be related to the different sero-prevalences in men and younger subjects, we compared these factors by sex and age group. The self-estimated health-related risk taking behaviour, sum of contacts and leisure-time activities decreased significantly by age in men and women (p<0.001; Appendix Table S4). The decrease in self-estimated health-related risk taking behaviour was most pronounced for age groups <35 yrs and 35-64 yrs, while number of contacts and leisure-time activities were substantially reduced in the oldest age group. Comparing men and women, self-estimated health-related risk taking behaviour was statistically higher for men than for women in the age group 35-65 yrs only (p<0.001, Appendix Table S5). In contrast, sum of contacts and number of leisure time activities was similar for men and women by age strata (p>0.05).

In order to check for effect modification by sex, age and behavioural factors, we calculated the sex- and age-stratified SARS-CoV-2 sero-incidence between baseline and follow-up by self-estimated health-related risk taking behaviour (Figure 8A), number of contacts outside own household (Figure 8B), number of leisure time activities in summer 2020 (Figure 8C). These data indicate a slightly higher risk of infection among men and women above the age of 34 years who indicated to have a high health-related risk taking behaviour compared to those with no high health-related risk taking behaviour. Men and women above the age of 64 years showed a higher SARS-CoV-2 sero-incidence if they had more personal contacts compared to participants having fewer personal contacts. Men with more leisure time activities in summer 2020 had a higher SARS-CoV-2 sero-incidence compared to less active men. However, all confidence intervals largely overlapped.

**Figure 8:**
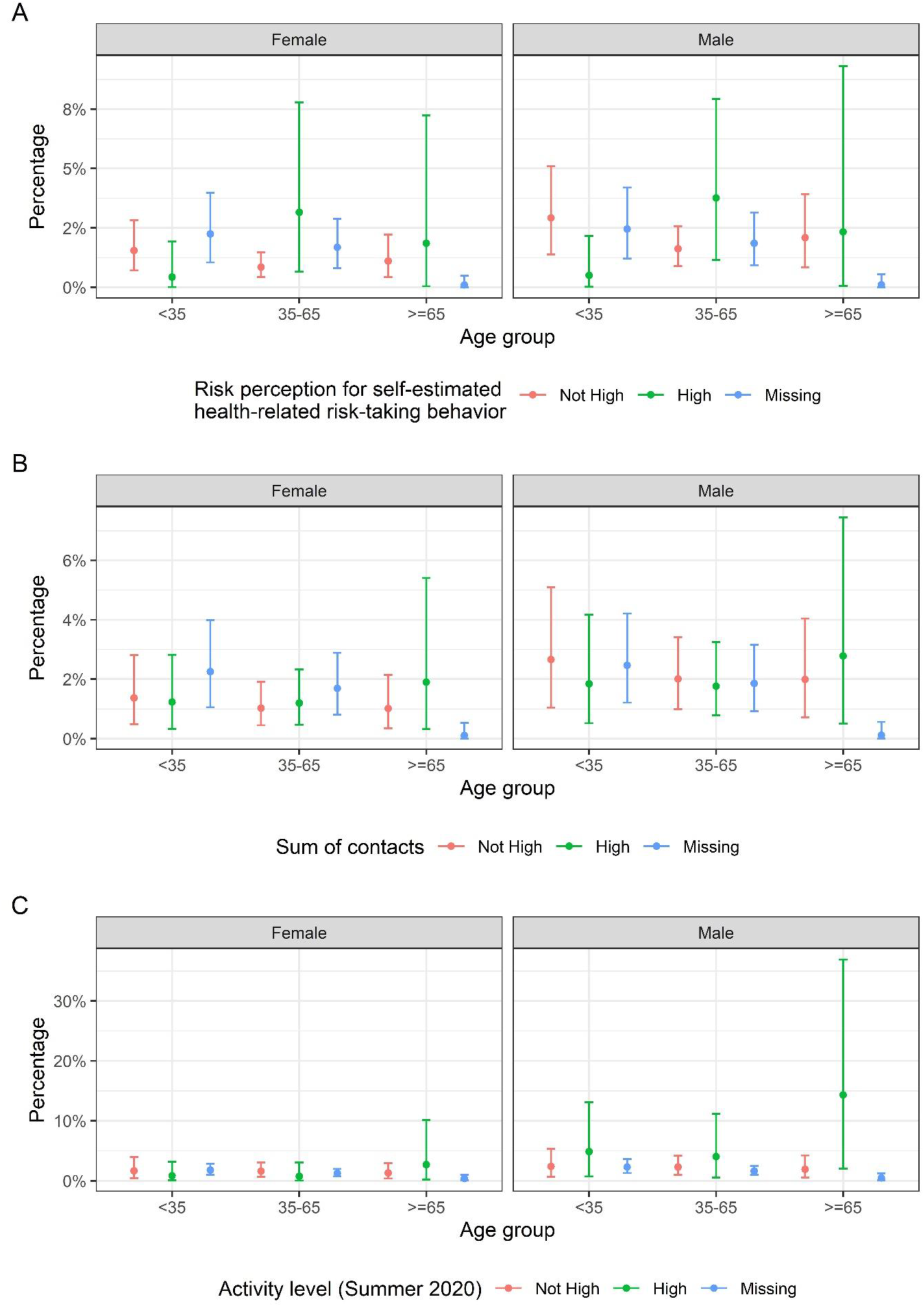
Sero-incidence of SARS-CoV-2 between baseline and follow-up by A) self-estimated health-related risk-taking behaviour, B) sum of contacts and C) leisure time activities in summer 2020 stratified for sex and age group.

## Discussion

Our data indicate a low SARS-CoV-2 sero-prevalence for the Munich general population living in private households eight months after the start of the pandemic. The incidence between the end of the first wave and the middle of the second wave was about as high as the related number of infections acquired during the first wave. Almost all sero-positive participants at baseline remained sero-positive at follow-up, indicating a high validity of the antibody test. Additionally, this supports previous reports suggesting that the humoral SARS-CoV-2 immune response is stable at least over the first eight months after infection (23, 24). We also showed a predominance in SARS-CoV-2 sero-positivity among male compared to female participants, and a reduced antibody prevalence with increasing age group.

Based on our data, the sero-prevalence for the Munich population above the age of 13 years living in private households was 3.6% (95% CI 2.9-4.3%). Until the end of November 2020, a total of 30,180 SARS-CoV-2 cases were officially registered in Munich (https://www.muenchen.de/rathaus/Stadtinfos/Coronavirus-Fallzahlen.html#Fallzahlen; Access date: 19-April-2021) which results in a population prevalence of 1.9%. This prevalence increased to 44,377 registered cases by the end of December 2020 (population prevalence 3.0%). The data are not directly comparable, as the official data also include children and persons living in institutions. While the prevalence of infection in children was at that time considered to be smaller than in adults, it was unknown for people living in institutions (e.g., homes for the elderly). Nevertheless, the comparison gives an indication that the percentage of officially registered infections improved considerably compared to the beginning of the pandemic. In a previous publication we estimated that solely one out of four infections was registered by the official infectious diseases surveillance system (3). A few population-based SARS-CoV-2 sero-studies have been conducted since the beginning of the pandemic (for review see (2)), most of them reporting sero-prevalences during or after the first wave. Up to now, only the Spanish national study reported the results of their follow-up data (25) with a sero-prevalence at follow-up (November 2020) of 5%, and thus comparable to our results.

In our study, one predictor of change in sero-prevalence from baseline to follow-up was male sex. While a higher risk of more severe COVID-19 among men was confirmed in several studies (26), findings on sex-differences in sero-prevalence are still inconsistent (2). As younger age was also related to a larger increase in SARS-CoV-2 sero-prevalence at follow-up, one might assume that differences in behaviour may contribute to these findings. We could confirm differences in health-risk taking behaviour, frequency of leisure-time activities, and number of contacts outside the own household especially by age. In the stratified analyses of the incidence of infection by age and sex, we observed a tendency that behaviour is related to higher sero-incidence of infection; although the low incidence and the reduced number of respondents to the questionnaires limited the statistical power of these analyses and result interpretation has to be done cautiously. However, the hypothesis that specific behaviour, i.e., restriction of contacts, might reduce the risk of infection is also supported by our observation that patients with autoimmune disease were at reduced risk of SARS-CoV-2 sero-positivity. This finding is in line with studies among, e.g., patients with inflammatory bowel disease (27). Overall, our findings support the notion that behavioural factors contribute to the spread of the pandemic, and therefore actions to increase adherence to public-health measures (such as information campaigns) are crucial especially in a time when acceptance of measures in the general population is faltering.

Our results also confirm the importance of household clustering while no indications for neighbourhood clustering were seen. The former finding is also supported by the observation that participants from higher income households were at non-significantly higher odds of SARS-CoV-2 sero-positivity. As we took into account total household income (not adjusted for number of persons in the household), single households were more likely to be in the lower income category and thus, at lower likelihood of household transmission.

We also saw a non-significant trend for lower odds of SARS-CoV-2 antibodies in smokers compared to non-smokers (OR 0.2; 95% CI 0.02-1.1), confirming results of a meta-analysis (28). Here, differences were mainly explained by differences in testing behaviour between smokers and non-smokers, which can be excluded in our study. One of the population-based studies published so far also indicated a lower SARS-CoV-2 sero-prevalence in smokers compared to non-smokers (29). Whether this is a true effect of, e.g., nicotine (30) or vitamin D (31) or result of some form of bias needs to be evaluated in future studies. Of note is also the tendency for higher odds of SARS-CoV-2 in participants pursuing school, however, the wide confidence interval does not permit strong conclusions.

Among the strengths of our study are its population-based, prospective nature in a large number of participants. Such population-based studies help authorities to plan public health measures based on the prevalence of exposure in the population, its spatial distribution and to further identify risk groups (32). With increasing availability of vaccines, this study design with further follow-ups will help public health authorities to understand the extent and duration of vaccine-induced immunity (33). We previously showed a high sensitivity and specificity of the Elecsys**®** Anti-SARS-CoV-2 assay (Roche) used in this sero-study (5). For the follow-up, we developed and carefully validated a semi-automated protocol using self-sampled DBS for SARS-CoV-2 serology (7). This approach facilitates field work to a very considerable extent and thus, makes studies with a higher frequency of follow-ups more feasible. Acceptance was high in our study population, and the percentage of participants lost to follow-up comparably low.

However, in the analyses we had to take selective participation into account by modelling the underlying non-response mechanism and calibrating the weights. This way, we could reduce attrition bias in our prevalence and incidence estimates. It is common in prospective cohort studies that baseline participants in younger age groups, with migration background and with lower socio-economic status are less likely to participate at follow-up (4). While typically participants with positive outcome are also more likely to participate in follow-up studies (34), our baseline participants who were SARS-CoV-2 antibody positive were less likely to take part at follow-up. This gives some indication that unknown sero-status motivated at least part of our baseline participants to take part in the study. Once positive sero-status was known to them, they might have lost interest. Further supported is this hypothesis by the fact that less participants were willing to complete the follow-up questionnaires than to take part in the SARS-CoV-2 antibody follow-up. As a consequence, statistical power to analyse the association between behavioural factor and SARS-CoV-2 sero-positivity was limited.

In conclusion, SARS-CoV-2 sero-prevalence in the Munich general population was still low by the end of 2020. Men and younger parts of the population were more likely to be affected. Risk-taking behaviour might be one reason for these differences. Therefore, non-pharmaceutical public health measures are still important.

## Data Availability

Data are accessible subject to data protection regulations upon reasonable request to the corresponding author. To facilitate reproducibility and reuse, the analysis and figure generation code has been made available on GitHub (https://github.com/koco19/epi2) and has been uploaded to ZENODO (https://doi.org/10.5281/zenodo.4707037) for long-term storage.

## Institutional Review Board Statement

The study was conducted in accordance with good clinical (GCP) and epidemiological practice (GEP) standards as well as the Declaration of Helsinki in its most recent form (as amended by the 64th WMA General Assembly, Fortaleza, Brazil, in October 2013). The study protocol was approved by the Institutional Review Board of the Medical Faculty at Ludwig Maximilian University Munich, Germany (opinion dated 31 March 2020; number 20–275; opinion date amendment: 10 October 2020), prior to study initiation.

## Informed Consent Statement

Informed consent was obtained from all study participants prior to study inclusion.

## Data Availability Statement

Our data are accessible to researchers upon reasonable request to the corresponding author taking data protection laws and privacy of study participants into account.

## Acknowledgments

We gratefully thank all study participants for their trust, time, data, and specimens. This study would also not have been possible without the staff of the Division of Infectious Diseases and Tropical Medicine at the University Hospital of LMU Munich, Helmholtz Centre Munich, and Bundeswehr Institute of Microbiology, as well as all medical students involved. We thank Judith Eckstein for outstanding support regarding public relations. We thank the teams from the press offices of LMU, University Hospital of LMU Munich, and of Helmholtz Centre Munich. We thank the KoCo19 advisory board members Stefan Endres, Stephanie Jacobs, Bernhard Liebl, Michael Mihatsch, Matthias Tschöp, Manfred Wildner, and Andreas Zapf. We thank Accenture for the development of the KoCo19 web-based survey application. We are grateful to the Statistical Office of the City of Munich, Germany, for providing statistical data on the Munich general population and to Landesamt für Gesundheit und Lebensmittelsicherheit for carrying out laboratory measurements. We also thank Guillaume Chauvet for advice on the sampling design and variance estimation and Dr. Joachim Heinrich for advice on inclusion of population density. We are grateful to the Munich police for their support in the fieldwork of the baseline study. The Munich Surgical Imaging GmbH, Cisco Systems, and the graphic/photo/IT infrastructure departments at the University Hospital of LMU Munich provided support during video production and online events. For fieldwork of the baseline study, BMW Group as part of their campaign “BMW hilft Helfenden” provided free cars. Mercedes-Benz Munich provided support with Mercedes-Benz Rent in the project infrastructure. MG acknowledges the support from the Joachim Herz Foundation through the Add-on Fellowship for Interdisciplinary Science.

## References

1. Rostami A, Sepidarkish M, Leeflang MMG, Riahi SM, Nourollahpour Shiadeh M, Esfandyari S, et al. SARS-CoV-2 seroprevalence worldwide: a systematic review and meta-analysis. Clin Microbiol Infect. 2021;27(3):331–40.

2. Lai CC, Wang JH, Hsueh PR. Population-based seroprevalence surveys of anti-SARS-CoV-2 antibody: An up-to-date review. Int J Infect Dis. 2020;101:314–22.

3. Pritsch M, Radon K, Bakuli A, Le Gleut R, Olbrich L, Guggenbuhl Noller JM, et al. Prevalence and Risk Factors of Infection in the Representative COVID-19 Cohort Munich. Int J Environ Res Public Health. 2021;18(3572).

4. Radon K, Saathoff E, Pritsch M, Guggenbuhl Noller JM, Kroidl I, Olbrich L, et al. Protocol of a population-based prospective COVID-19 cohort study Munich, Germany (KoCo19). BMC Public Health. 2020;20(1):1036.

5. Olbrich L, Castelletti N, Schälte Y, Garí M, Pütz P, Bakuli A, et al. A Serology Strategy for Epidemiological Studies Based on the Comparison of the Performance of Seven Different Test Systems - The Representative COVID-19 Cohort Munich. medRxiv. 2021.

6. Iglesias C, Torgerson D. Does length of questionnaire matter? A randomised trial of response rates to a mailed questionnaire. J Health Serv Res Policy. 2000;5(4):219–21.

7. Beyerl J, Rubio-Acero R, Castelletti N, Panovic I, Kroidl I, Khan ZN, et al. A Dried Blood Spot Protocol for high throughput analysis of SARS-CoV-2 Serology based on the Roche Elecsys Anti-N Assay. Submitted. 2021.

8. Sempos CT, Tian L. Adjusting Coronavirus Prevalence Estimates for Laboratory Test Kit Error. Am J Epidemiol. 2020.

9. Särndal CE, Swensson B, Wretman J. Model Assisted Survey Sampling. New York: Springer; 2003.

10. Deville JC, Särndal CE. Calibration estimators in survey sampling. J Am Stat Assoc. 1992;87(418):376–82.

11. Deville JC. Variance estimation for complex statistics and estimators: linearization and residual techniques.. Surv Meth. 1999;25(2):193–204.

12. Juillard H, Chauvet G. Variance estimation under monotone non-response for a panel survey. Surv Meth. 2018;44(2).

13. Cameron AC, Gelbach JB, Miller DL. Bootstrap-based improvements for inference with clustered errors. Rev Econ Stat. 2008;90(3):414–27.

14. Nichols TE, Holmes AP. Nonparametric permutation tests for functional neuroimaging: a primer with examples. Hum Brain Mapp. 2002;15(1):1–25.

15. Lee D. CARBayes: An R Package for Bayesian Spatial Modeling with Conditional Autoregressive Priors. J Stat Software. 2013;55(13):1–24.

16. Leroux B, Lei X, Breslow N. Estimation of Disease Rates in Small Areas: A New Mixed Model for Spatial Dependence. In: Halloran ME, Berry D, editors. Models in Epidemiology, the Environment and Clinical Trials. New York: Springer; 1999. p. 135–78.

17. Lee D, Mitchell R. Boundary detection in disease mapping studies. Biostatistics. 2012;13(3):415–26.

18. Lee D. A comparison of conditional autoregressive models used in Bayesian disease mapping. Spat Spatiotemporal Epidemiol. 2011;2(2):79–89.

19. Bürkner P. Advances Bayesian multilevel modeling with the R package brms. R Journal. 2018;10(1):395–411.

20. Bürkner P. brms: an R package for Bayesian multilevel models using stan J Stat Software. 2017;80(1):1–28.

21. Erler NS, Rizopoulos D, Lesaffre EM. JointAI: Joint Analysis and Imputation of Incomplete Data in R. J Stat Software. 2020; in press.

22. Erler NS, Rizopoulos D, Rosmalen J, Jaddoe VW, Franco OH, Lesaffre EM. Dealing with missing covariates in epidemiologic studies: a comparison between multiple imputation and a full Bayesian approach. Stat Med. 2016;35(17):2955–74.

23. Dan JM, Mateus J, Kato Y, Hastie KM, Yu ED, Faliti CE, et al. Immunological memory to SARS-CoV-2 assessed for up to 8 months after infection. Science. 2021;371(6529).

24. He Z, Ren L, Yang J, Guo L, Feng L, Ma C, et al. Seroprevalence and humoral immune durability of anti-SARS-CoV-2 antibodies in Wuhan, China: a longitudinal, population-level, cross-sectional study. Lancet. 2021;397(10279):1075–84.

25. Pérez-Olmeda M, Saugar JM, Fernández-García A, Pérez-Gómez B, Pollán M, Avellón A, et al. Evolution of antibodies against SARS-CoV-2 over seven months: experience of the Nationwide Seroprevalence ENE-COVID Study in Spain. medRxiv. 2021.

26. Takahashi T, Ellingson MK, Wong P, Israelow B, Lucas C, Klein J, et al. Sex differences in immune responses that underlie COVID-19 disease outcomes. Nature. 2020;588(7837):315–20.

27. Stallmach A, Sturm A, Blumenstein I, Helwig U, Koletzko S, Lynen P, et al. Addendum to S3-Guidelines Crohn’s disease and ulcerative colitis: Management of Patients with Inflammatory Bowel Disease in the COVID-19 Pandemic - open questions and answers. Z Gastroenterol. 2020;58(10):982–1002.

28. Simons D, Shahab L, Brown J, Perski O. The association of smoking status with SARS-CoV-2 infection, hospitalization and mortality from COVID-19: a living rapid evidence review with Bayesian meta-analyses (version 7). Addiction. 2020.

29. Carrat F, de Lamballerie X, Rahib D, Blanché H, Lapidus N, Artaud F, et al. Seroprevalence of SARS-CoV-2 among adults in three regions of France following the lockdown and associated risk factors: a multicohort study. medRxiv. 2020.

30. Farsalinos K, Niaura R, Le Houezec J, Barbouni A, Tsatsakis A, Kouretas D, et al. Editorial: Nicotine and SARS-CoV-2: COVID-19 may be a disease of the nicotinic cholinergic system. Toxicol Rep. 2020;7:658–63.

31. Khan AH, Nasir N, Nasir N, Maha Q, Rehman R. Vitamin D and COVID-19: is there a role? J Diabetes Metab Disord. 2021:1–8.

32. Peeling RW, Olliaro PL. The time to do serosurveys for COVID-19 is now. Lancet Respir Med. 2020;8(9):836–8.

33. Eckerle I, Meyer B. SARS-CoV-2 seroprevalence in COVID-19 hotspots. Lancet. 2020;396(10250):514–5.

34. Forster F, Kreissl S, Wengenroth L, Vogelberg C, von Mutius E, Schaub B, et al. Third Follow-Up of the Study on Occupational Allergy Risks (SOLAR III) in Germany: Design, Methods, and Initial Data Analysis. Front Public Health. 2021;9:591717.

