## Appendix for "From first to second wave: follow-up of the prospective Covid-19 cohort (KoCo19) in Munich (Germany)"

### 1. Analysis of sero-prevalence over time in our study

We analysed the sero-positivity over time using the aggregated proportion of the sero-positive results over the time intervals. We fitted a count data regression to estimate the counts of sero-positive samples using a one-way ANOVA model within the Bayesian modelling using non-informative priors for the individual weeks as normal distribution with mean zero and standard deviation 10. The estimates were based on 5,000 warmup MCMC samples followed by another 5,000 MCMC samples used for the posterior distribution (1, 2). For the count data regression, we could aggregate two options, one without the sampling weights. We explored the outcomes as binomial counts versus Poisson counts for sensitivity analysis. However, the results remain unchanged and we could observe an increase in sero-prevalence in the population during the later time points. In addition to this, as described in Figure 2, dried blood samples (DBS) of individuals (including household members) who produced intermediate results or positive results retested using venous blood samples. We performed the venous blood draw only during the last time point of the study, and hence the increase in the sero-positivity at the last week most likely have artificial effect. Thus, we compared the results of sero-positivity only using the DBS versus the results using the combination of DBS and reconfirmation with venous blood draw. The increase during the last two time points would be obvious, however it is higher in the last week of the study when we had the additional results using venous blood draw. The DBS results and the venous blood results are in Table S1 and the sero-positivity over time is illustrated in Figure S1 and Figure S2.

**Table S1: Comparison of the follow-up plasma results, using DBS or Venous Blood Samples**

| Venous Blood Sample | Dried Blood Sample Results | | | |
|---|---|---|---|---|
| | Intermediate | Negative | Positive | Total |
| Negative | 11 (12.0%) | 76 (82.6%) | 5 ( 5.4%) | 92 (100.0%) |
| Positive | 12 (13.5%) | 3 ( 3.4%) | 74 (83.1%) | 89 (100.0%) |
| Not Done | 0 ( 0.0%) | 4208 (99.0%) | 44 ( 1.0%) | 4252 (100.0%) |
| Total | 23 ( 0.5%) | 4287 (96.7%) | 123 ( 2.8%) | 4433 (100.0%) |

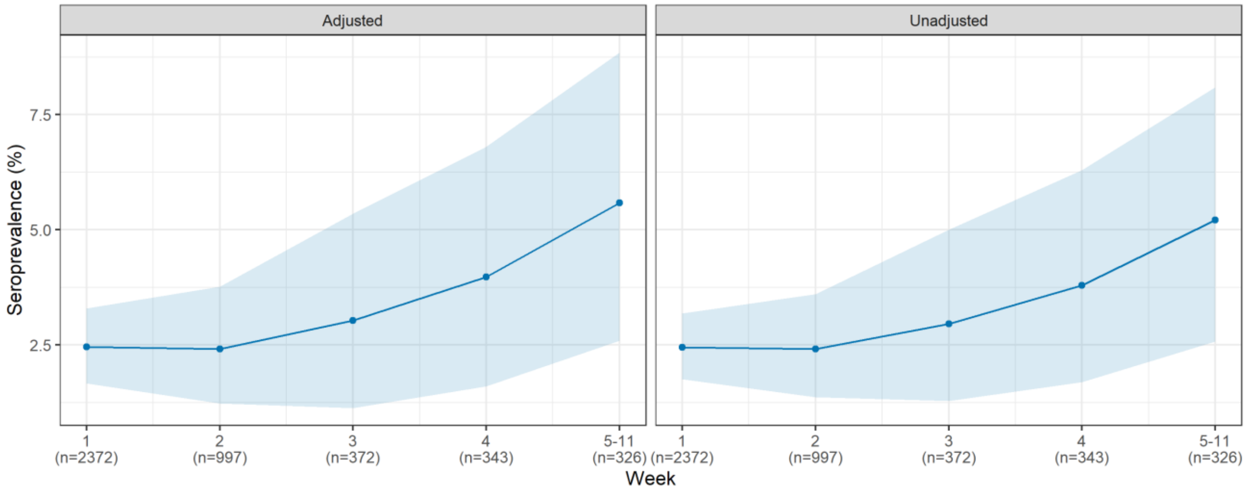


**Figure S1: For sensitivity and specificity adjusted (left) and unadjusted (right) SARS-CoV-2 sero-prevalence over the follow-up period excluding DBS intermediates. The 95% confidence intervals for the weekly sero-prevalence are based on the 2.5 and 97.5 percentiles from 5,000 repetitions of a cluster bootstrap that accounts for within household clustering. The estimates do not account for sample weights.**

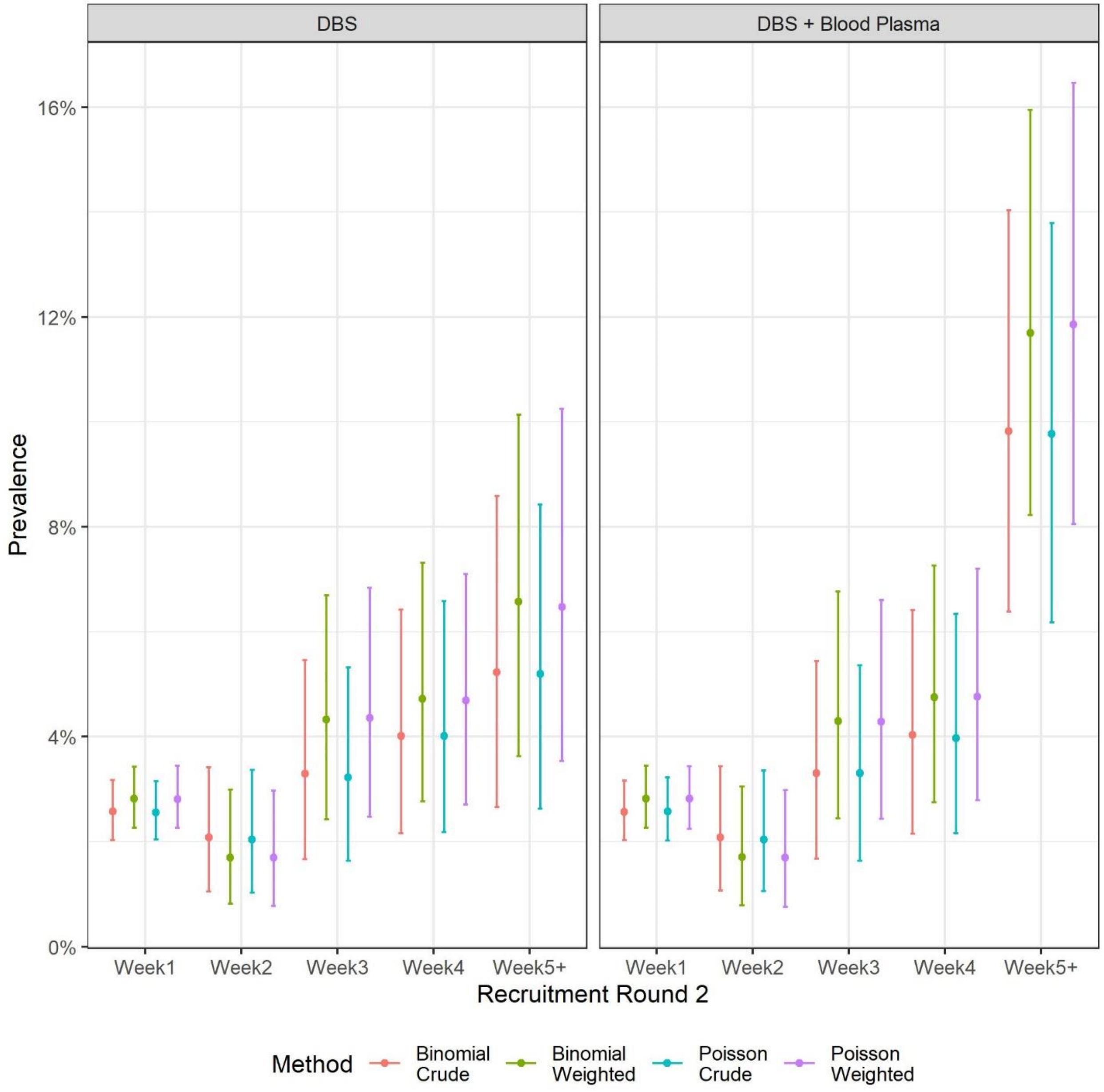


**Figure S2: Comparison of the sero-positivity over the follow-up period accounting for sampling weights.**

### 2. Analysis of sero-prevalence over the spatial structure of Munich

We estimated the sero-prevalence over the set of non-overlapping spatial areal units, which are the 25 districts or boroughs of Munich. Our goal was to test the hypothesis of the association in sero-prevalence and the population density of the boroughs. We would expect the observations of sero-prevalence from boroughs closer together tending to have similar values. We report the spatial autocorrelation present in the prevalence observed in the different boroughs using the Moran's I. We report the Moran's I permutation test for spatial autocorrelation based on 10,000 random permutations, using the functionality of the spatial weights (3, 4) ☐We considered two kinds of spatial weighting schemes, starting from a binary neighbours list, in which boroughs are either listed as neighbours or are absent (thus not in the set of neighbours for some definition), and row standardised Euclidian distances from a borough (sums over all links to each borough). Using both schemes we did not observe any spatial autocorrelation with the Moran's I being 0.025 and 0.015 respectively and a corresponding p-value of 0.271 and 0.303. This allowed us not to reject the null hypothesis that the crude borough- wide sero-prevalence estimates had a spatial autocorrelation significantly different from zero. However, in case of population density within the boroughs the Moran's I was computed to be 0.291 and 0.317 respectively with a p-value of 0.007 and 0.005. Thus, there is evidence for spatial autocorrelation in the population density estimates.

Since the population density estimates are spatially correlated, when evaluating the association between population density and sero-prevalence at the level of boroughs within Munich, we used the common remedy for this spatial autocorrelation by augmenting the linear predictor with a set of spatially correlated random effects, as part of a Bayesian hierarchical model (3, 4) ☐The random effects are considered through a conditional autoregressive model (CAR), which induces spatial autocorrelation through the adjacency structure of the boroughs (spatial units within the city of Munich). Among the CAR priors that are used in practice, we explored the two options, one using the global and the second using a local CAR prior. A binary specification based on geographical neighbourhood contact is used, where $w_{kj} = 1$ if boroughs $\left(S_k, S_j\right)$ share a common border (denoted $k \sim j$), and is zero otherwise. This specification forces the outcomes of geographically adjacent or neighbouring boroughs ($w_{kj} = 1$) to be correlated, and random effects related to non-neighbouring boroughs to be conditionally independent given the values of the remaining random effects. We used the CAR model as was proposed by Leroux, Lei and Breslow (1999) where one random effect is used for modelling the differential intensity of the spatial autocorrelation for the globally smoothed spatial sero-prevalence rates (3–5). However, it has been argued often that in more complex urban setup (that would be typical for a city like Munich); there might be possibilities for a localized spatial structure. In Munich city too, there exist spatial pockets of super high density surrounded by spatial pockets of lower population density. The model proposed by Lee and Mitchell (2012) considered the partial correlation between random effects in adjacent spatial units as a function of their dissimilarity (3, 4, 6, 7). So we considered not only a binary geographical neighbourhood but also an additional component of the density difference between two neighbouring boroughs (rescaled by the standard deviation among all neighbours) to create a dissimilarity matrix for the boroughs. This allowed for local CAR prior models. Both kind of models were done to estimate the counts of sero-positives in a borough at an aggregated level, both using the sample weights and ignoring them as potentially binomial or Poisson counts. The estimates were based on 10,000 burn in MCMC samples followed by another 15,000 MCMC samples used for the posterior distribution. Additionally adjustment by population density based counts could be obtained from such models. This allowed us to have sensitivity analyses of the sero-prevalence across the boroughs within Munich (Figure S3 – Globally smoothed CAR model and S4 Local smoothed CAR model). We observed that sero-prevalence estimates varied slightly across the boroughs however, the confidence intervals were majorly overlapping.

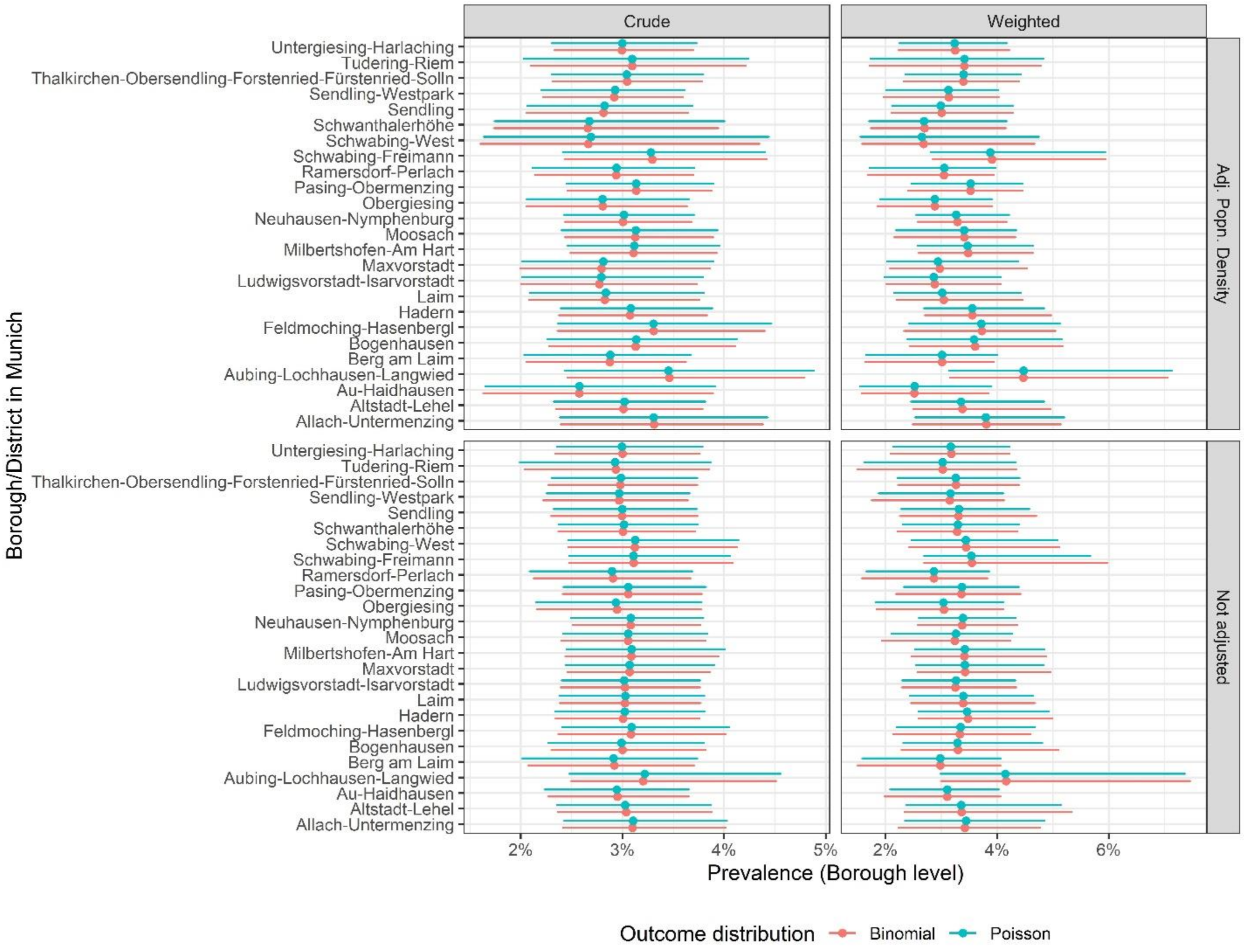


**Figure S3: Sero-prevalence estimates across the different boroughs of Munich using CAR model priors with a single level of spatial autocorrelation as random effects.**

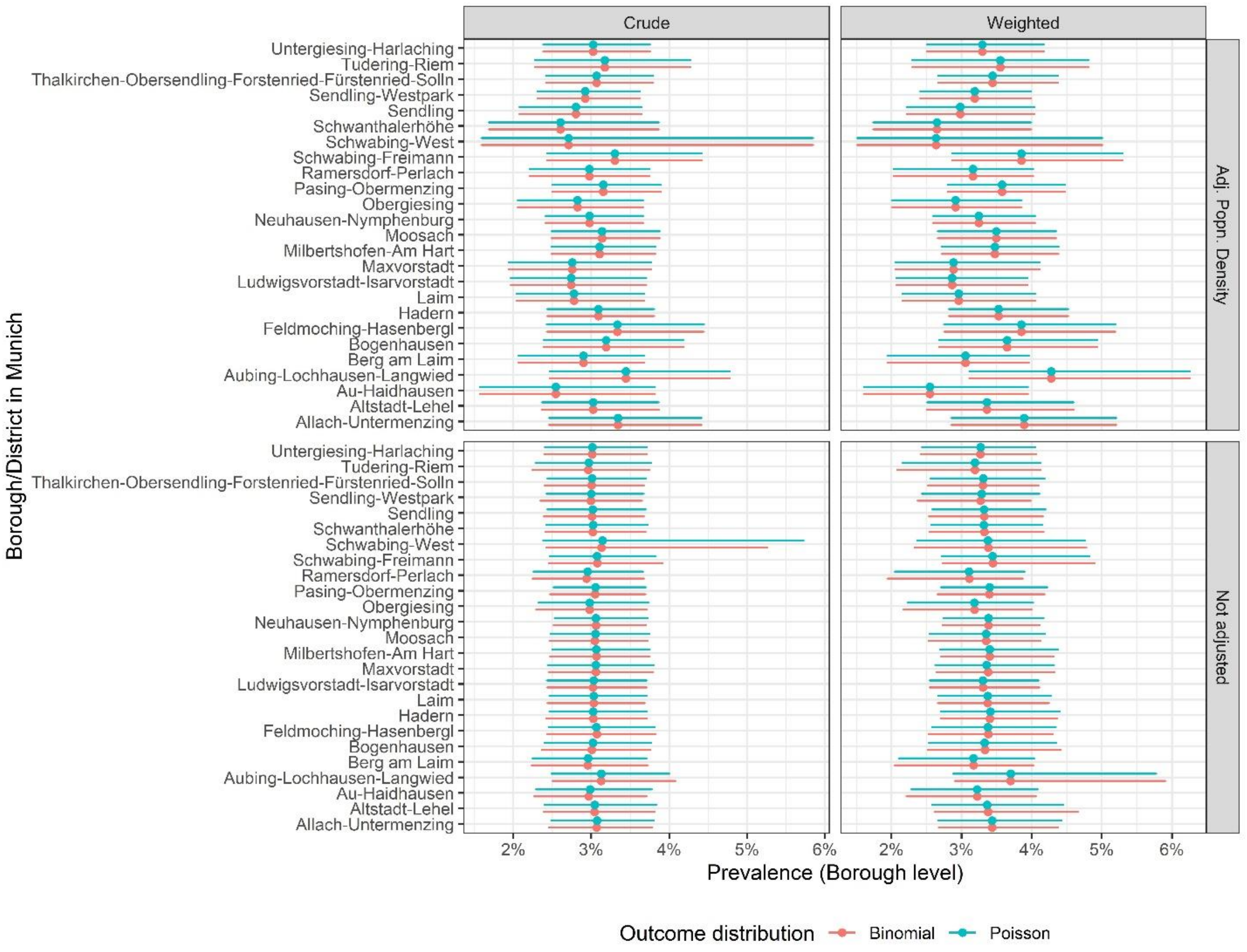


**Figure S4 Sero-prevalence estimates across the different boroughs of Munich with CAR model priors through the dissimilarity of the population density in neighbouring boroughs for spatial autocorrelation.**

In the above analyses we observed that the population density of the boroughs are not likely to be associated with sero-prevalence estimates. Adjusted sero-prevalence estimates changed slightly. However, the confidence intervals were overlapping. Nevertheless, this analysis was performed at the aggregated level of boroughs. In reality, boroughs are not completely homogenous population units within the city of Munich. Thus, constituencies within a borough offer a higher level of homogeneity of population density than the borough itself. In our representative sample, we have data from only 361 of the 755 constituencies in Munich. So performing a spatial analysis at the level of the constituencies was difficult, since it is likely that we often missed data from neighbouring constituencies. We additionally investigated into the association of population density and sero-prevalence at the aggregation level of the constituency. Another difficulty in the measurement of the association is that only 89 of the 361 sampled constituencies had any sero-positive counts. Thus, there exists an excess of zeros in the positivity count. To tackle the issue of more zeros than would be in a Poisson outcome, we considered the Poisson distribution to obtain the estimated counts of positive individuals at the level of constituency with an additional adjustment for the zero counts. The zero inflated models are two-component mixture models combining a point mass at zero with a count distribution. There exists two sources of zeros: zeros may come from both the point mass and from the count component. For modelling the unobserved state (zero vs. count), a binary model, and a separate count component is used (8). We also compared the estimates using a negative binomial outcome. However, the results remained unchanged and no statistically significant association between the zero component or the count component and the population density of the constituency was observed (Table S2).

**Table S2 Comparison of the estimates for the covariate of population density at the constituency level using the Poisson and the negative binomial model.**

| **Model** | **Zero Inflated Poisson** | | | **Zero Inflated Negative Binomial** | | |
|---|---|---|---|---|---|---|
| **Covariate** | Estimate | 2.50% | 97.50% | Estimate | 2.50% | 97.50% |
| **Count (Intercept)** | -2.468 | -2.810 | -2.126 | -2.963 | -3.952 | -1.973 |
| **Count Population Density March 2020** | -0.002 | -0.005 | 0.001 | -0.001 | -0.006 | 0.003 |
| **Zero (Intercept)** | 0.394 | -0.270 | 1.058 | -0.806 | -4.022 | 2.411 |
| **Zero Population Density March 2020** | -0.003 | -0.009 | 0.004 | -0.007 | -0.029 | 0.015 |

### Additional tables and figures

**Table S3: Course of SARS-CoV-2 antibody status within the KoCo19 follow-up participants**

| | | | **SARS-CoV-2...** | **...sero-remission** (positive at baseline, negative at follow-up) | | **...sero-persistence** (positive at baseline and follow-up) | | **...sero-incidence** (negative at baseline, positive at follow-up) | | | **...seroprevalence** (ever positive) | | |
|---|---|---|---|---|---|---|---|---|---|---|---|---|---|
| **Variable** | **Categories*** | **N** | **$n_{Missing}$** | **n** | **%** | **n** | **%** | **n** | **%** | **p** | **n** | **%** | **p** |
| | **Total** | **4433** | **0** | **6** | **0.14** | **64** | **1.44** | **71** | **1.60** | | **141** | **3.18** | |
| **Sex** | Female | 2320 | 0 | 5 | 0.22 | 30 | 1.29 | 32 | 1.38 | 0.20 | 67 | 2.89 | 0.30 |
| | Male | 2113 | | 1 | 0.05 | 34 | 1.61 | 39 | 1.85 | | 74 | 3.50 | |
| **Age** | 0-19 | 212 | 0 | 0 | 0.00 | 3 | 1.42 | 6 | 2.83 | 0.86 | 9 | 4.25 | 0.70 |
| (years) | 20-34 | 1040 | | 0 | 0.00 | 16 | 1.54 | 18 | 1.73 | | 34 | 3.27 | |
| | 35-49 | 1271 | | 3 | 0.24 | 19 | 1.49 | 22 | 1.73 | | 44 | 3.46 | |
| | 50-64 | 1166 | | 2 | 0.17 | 14 | 1.20 | 18 | 1.54 | | 34 | 2.92 | |
| | 65-79 | 599 | | 1 | 0.17 | 11 | 1.84 | 6 | 1.00 | | 18 | 3.01 | |
| | 80+ | 145 | | 0 | 0.00 | 1 | 0.69 | 1 | 0.69 | | 2 | 1.38 | |
| **Birth country** | Germany | 3521 | 232 | 4 | 0.11 | 49 | 1.39 | 53 | 1.51 | 0.59 | 106 | 3.01 | 0.34 |
| | Other | 680 | | 2 | 0.29 | 10 | 1.47 | 13 | 1.91 | | 25 | 3.68 | |
| **Level of education** | Student | 80 | 446 | 0 | 0.00 | 1 | 1.25 | 4 | 5.00 | 0.06 | 5 | 6.25 | 0.2 |
| | <12 yrs | 1175 | | 2 | 0.17 | 17 | 1.45 | 12 | 1.02 | | 33 | 2.81 | |
| | ≥12 yrs | 2732 | | 4 | 0.15 | 40 | 1.46 | 47 | 1.72 | | 89 | 3.26 | |
| **Occupationally active** | No | 784 | 236 | 0 | 0.00 | 6 | 0.77 | 8 | 1.02 | 0.09 | 14 | 1.79 | 0.02 |
| | Yes | 3413 | | 6 | 0.18 | 53 | 1.55 | 58 | 1.70 | | 117 | 3.43 | |
| **Smoking status** | Never smoker | 2217 | 253 | 3 | 0.14 | 29 | 1.31 | 41 | 1.85 | 0.66 | 73 | 3.29 | 0.55 |
| | Ex-smoker | 1234 | | 2 | 0.16 | 19 | 1.54 | 19 | 1.54 | | 40 | 3.24 | |
| | Current smoker | 729 | | 1 | 0.14 | 11 | 1.51 | 6 | 0.82 | | 18 | 2.47 | |

| | | | SARS-CoV-2... | ...sero-remission (positive at baseline, negative at follow-up) | | ...sero-persistence (positive at baseline and follow-up) | | ...sero-incidence (negative at baseline, positive at follow-up) | | | ...seroprevalence (ever positive) | | |
|---|---|---|---|---|---|---|---|---|---|---|---|---|---|
| **General health** | Excellent | 686 | 233 | 0 | 0.00 | 10 | 1.46 | 15 | 2.19 | 0.84 | 25 | 3.64 | 0.72 |
| | Very good | 1852 | | 3 | 0.16 | 26 | 1.40 | 30 | 1.62 | | 59 | 3.19 | |
| | Good | 1493 | | 3 | 0.20 | 20 | 1.34 | 18 | 1.21 | | 41 | 2.75 | |
| | Not good | 169 | | 0 | 0.00 | 3 | 1.78 | 2 | 1.18 | | 5 | 2.96 | |
| **Household type** | Single | 588 | 254 | 0 | 0.00 | 11 | 1.87 | 6 | 1.02 | 0.66 | 17 | 2.89 | 0.28 |
| | Couple | 1531 | | 2 | 0.13 | 18 | 1.18 | 20 | 1.31 | | 40 | 2.61 | |
| | Family | 1674 | | 3 | 0.18 | 24 | 1.43 | 29 | 1.73 | | 56 | 3.35 | |
| | Others | 386 | | 0 | 0.00 | 8 | 2.07 | 8 | 2.07 | | 17 | 4.40 | |
| **Household income** | ≤2500 | 501 | 1210 | 1 | 0.20 | 6 | 1.20 | 4 | 0.80 | 0.2 | 11 | 2.20 | 0.02 |
| (Euro) | 2501-4000 | 706 | | 1 | 0.14 | 6 | 0.85 | 8 | 1.13 | | 15 | 2.12 | |
| | 4001-6000 | 1043 | | 1 | 0.10 | 13 | 1.25 | 18 | 1.73 | | 32 | 3.07 | |
| | 6000+ | 973 | | 3 | 0.31 | 15 | 1.54 | 26 | 2.67 | | 44 | 4.52 | |
| **Living area/inhabitant** | ≤ 30 | 1432 | 269 | 2 | 0.14 | 20 | 1.40 | 22 | 1.54 | 0.2 | 44 | 3.07 | 0.03 |
| (sqm/individual) | 31-40 | 1038 | | 1 | 0.10 | 19 | 1.83 | 23 | 2.22 | | 43 | 4.14 | |
| | 41-55 | 889 | | 2 | 0.22 | 7 | 0.79 | 7 | 0.79 | | 16 | 1.80 | |
| | 55+ | 805 | | 1 | 0.12 | 15 | 1.86 | 11 | 1.37 | | 27 | 3.35 | |
| **Building type** | 1-2 apt | 1263 | 0 | 2 | 0.16 | 19 | 1.50 | 19 | 1.50 | 0.37 | 40 | 3.17 | 0.11 |
| (No of apartments) | 3-4 apt | 307 | | 1 | 0.33 | 2 | 0.65 | 0 | 0.00 | | 3 | 0.98 | |
| | 5+ apt | 2856 | | 3 | 0.11 | 43 | 1.51 | 52 | 1.82 | | 98 | 3.43 | |
| | Others | 7 | | 0 | 0.00 | 0 | 0.00 | 0 | 0.00 | | 0 | 0.00 | |
| **Health-risk taking** | Not high | 2156 | 1668 | 4 | 0.19 | 29 | 1.35 | 33 | 1.53 | 0.63 | 66 | 3.06 | 0.6 |
| **behavior** | High | 609 | | 0 | 0.00 | 11 | 1.81 | 10 | 1.64 | | 21 | 3.45 | |
| **Leisure time activity level** | | | | | | | | | | | | | |

| | | | SARS-CoV-2... | ...sero-remission (positive at baseline, negative at follow-up) | | ...sero-persistence (positive at baseline and follow-up) | | ...sero-incidence (negative at baseline, positive at follow-up) | | | ...seroprevalence (ever positive) | | |
|---|---|---|---|---|---|---|---|---|---|---|---|---|---|
| After 1st wave | Not high | 1052 | 3170 | 1 | 0.10 | 13 | 1.24 | 19 | 1.81 | 0.75 | 33 | 3.14 | 0.67 |
| (June-Oct 2020) | High | 211 | | 0 | 0.00 | 2 | 0.95 | 6 | 2.84 | | 8 | 3.79 | |
| **Sum of personal contacts** | | Mean (SD) | | Mean | SD | Mean | SD | Mean | SD | | Mean | SD | |
| After 1st wave (June-Oct 2020) | SARS-CoV-2 negative | 8.81 (4.28) | 1665 | 11.5 | 44.382.00 | 10.7 | 3.89 | 8.91 | 4.80 | 0.02 | 9.85 / 8.81 | 4.46 / 4.28 | 0.03 |

*for category definitions please see methods section of the article*

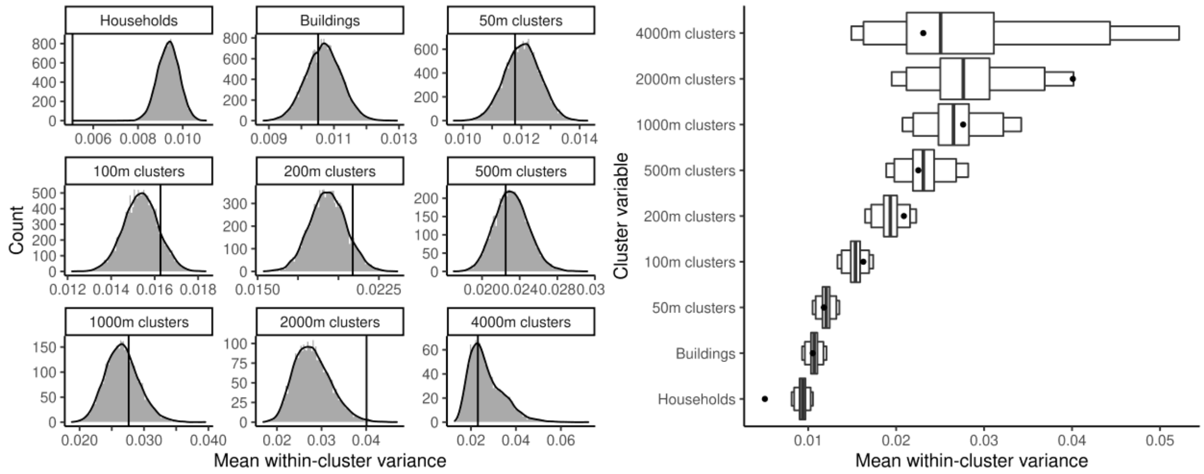


**Figure S5: Distribution of mean within-cluster variance of test results under 10,000 random permutation of cluster assignments, with clusters being households, buildings, and geospatial clusters of different sizes. Household membership left invariant for building and geospatial clusters. Left: Value distribution. Right: 50%, 95%, 99% CIs. Black lines (left) and dots (right) indicate the observed values.**

**Table S4: Summary across age group and sex for the response to the behaviour questionnaires stratified by sex. p- values are from Pearson's Chi-squared test with simulated p-value (based on 10000 replicates).**

| | | **Age group** | | | |
|---|---|---|---|---|---|
| **Sex** | | <35 (N=1252) | 35-65 (N=2437) | >=65 (N=744) | p value |
| | | Health-risk taking Behaviour | | | < 0.001 |
| Female | High | 66 (17.2%) | 75 (8.9%) | 27 (9.5%) | |
| | Not High | 317 (82.8%) | 766 (91.1%) | 258 (90.5%) | |
| | Missing | 272 | 425 | 114 | |
| | | Sum of contacts | | | < 0.001 |
| | High | 176 (45.8%) | 360 (42.8%) | 51 (17.9%) | |
| | Not High | 208 (54.2%) | 482 (57.2%) | 234 (82.1%) | |
| | Missing | 271 | 424 | 114 | |
| | | Leisure time activity level | | | < 0.001 |
| | Not High | 105 (64.4%) | 299 (87.4%) | 168 (88.9%) | |
| | High | 58 (35.6%) | 43 (12.6%) | 21 (11.1%) | |
| | Missing | 492 | 924 | 210 | |
| | | Health-risk taking Behaviour | | | < 0.001 |
| Male | High | 62 (21.8%) | 106 (14.1%) | 22 (10.0%) | |
| | Not High | 222 (78.2%) | 646 (85.9%) | 198 (90.0%) | |
| | Missing | 313 | 419 | 125 | |
| | | Sum of contacts | | | < 0.001 |
| | High | 121 (42.6%) | 359 (47.7%) | 50 (22.6%) | |
| | Not High | 163 (57.4%) | 393 (52.3%) | 171 (77.4%) | |
| | Missing | 313 | 419 | 124 | |
| | | Leisure time activity level | | | < 0.001 |
| | Not High | 101 (73.7%) | 258 (86.0%) | 121 (91.7%) | |
| | High | 36 (26.3%) | 42 (14.0%) | 11 (8.3%) | |
| | Missing | 460 | 871 | 213 | |

**Table S5: Summary across age group and sex for the response to the behaviour questionnaires stratified by age group. p- values are from Pearson's Chi-squared test with simulated p-value (based on 10000 replicates).**

| | | Sex | | |
|---|---|---|---|---|
| **Age group** | | Female (N=2320) | Male (N=2113) | p value |
| | | Health-risk taking Behaviour | | 0.137 |
| | High | 66 (17.2%) | 62 (21.8%) | |
| | Not High | 317 (82.8%) | 222 (78.2%) | |
| | Missing | 272 | 313 | |
| | | Sum of contacts | | 0.431 |
| | High | 176 (45.8%) | 121 (42.6%) | |
| <35 | Not High | 208 (54.2%) | 163 (57.4%) | |
| | Missing | 271 | 313 | |
| | | Leisure time activity level | | 0.104 |
| | Not High | 105 (64.4%) | 101 (73.7%) | |
| | High | 58 (35.6%) | 36 (26.3%) | |
| | Missing | 492 | 460 | |
| | | Health-risk taking Behaviour | | 0.001 |
| | High | 75 (8.9%) | 106 (14.1%) | |
| | Not High | 766 (91.1%) | 646 (85.9%) | |
| | Missing | 425 | 419 | |
| | | Sum of contacts | | 0.049 |
| | High | 360 (42.8%) | 359 (47.7%) | |
| 35-65 | Not High | 482 (57.2%) | 393 (52.3%) | |
| | Missing | 424 | 419 | |
| | | Leisure time activity level | | 0.641 |
| | Not High | 299 (87.4%) | 258 (86.0%) | |
| | High | 43 (12.6%) | 42 (14.0%) | |
| | Missing | 924 | 871 | |
| | | Health-risk taking Behaviour | | 0.88 |
| | High | 27 (9.5%) | 22 (10.0%) | |
| | Not High | 258 (90.5%) | 198 (90.0%) | |
| >=65 | Missing | 114 | 125 | |
| | | Sum of contacts | | 0.217 |
| | High | 51 (17.9%) | 50 (22.6%) | |
| | Not High | 234 (82.1%) | 171 (77.4%) | |

| Age group | | Sex | | |
|---|---|---|---|---|
| | | Female (N=2320) | Male (N=2113) | p value |
| | Missing | 114 | 124 | |
| | | Leisure time activity level | | 0.454 |
| | Not High | 168 (88.9%) | 121 (91.7%) | |
| | High | 21 (11.1%) | 11 (8.3%) | |
| | Missing | 210 | 213 | |

### References


1. Bürkner P. Advances Bayesian multilevel modeling with the R package brms. R Journal. 2018;10(1):395-411.
2. Bürkner P. brms: an R package for Bayesian multilevel models using stan J Stat Software. 2017;80(1):1-28.
3. Lee D. CARBayes: An R Package for Bayesian Spatial Modeling with Conditional Autoregressive Priors. J Stat Software. 2013;55(13):1-24.
4. Leroux B, Lei X, Breslow N. Estimation of Disease Rates in Small Areas: A New Mixed Model for Spatial Dependence. In: Halloran ME, Berry D, editors. Models in Epidemiology, the Environment and Clinical Trials. New York: Springer; 1999. p. 135-78.
5. Lee D, Mitchell R. Boundary detection in disease mapping studies. Biostatistics. 2012;13(3):415-26.
6. Lee D. A comparison of conditional autoregressive models used in Bayesian disease mapping. Spat Spatiotemporal Epidemiol. 2011;2(2):79-89.
7. Zeileis A, Kleiber C, Jackman S. Regression models for count data in R. J Stat Software. 2008;27(8):1-25.
8. Lambert D. Zero-inflated Poisson regression, with an application to defects in manufacturing. Technometrics. 1992;34(1):1-14.
